# Proton pump inhibitor use is not associated with severe COVID-19 related outcomes: A propensity score weighted analysis of a national veteran cohort

**DOI:** 10.1101/2021.07.22.21258785

**Authors:** Shailja C. Shah, Alese E. Halvorson, Brandon McBay, Chad Dorn, Otis Wilson, Jason Denton, Sony Tuteja, Kyong-Mi Chang, Kelly Cho, Richard L. Hauger, Ayako Suzuki, Christine M. Hunt, Edward Siew, Michael E. Matheny, Adriana Hung, Robert A. Greevy, Christianne L. Roumie

## Abstract

**Background and Aims:** Low pH deactivates most pathogens, including coronaviruses. Proton pump inhibitors (PPIs) are potent gastric acid suppressing medications. Whether PPI use vs non-use is associated with severe Coronavirus disease-2019 (COVID-19) outcomes remains uncertain. We aimed to compare severe COVID-19 outcomes between current outpatient PPI users and non-users.

**Methods:** We conducted a retrospective propensity score-weighted analysis of a national cohort of US veterans with established care who tested positive for severe acute respiratory syndrome coronavirus-2 (SARS-CoV-2) through January 9, 2021, and who had 60 days of follow-up. The positive test date was the index date. Current outpatient PPI use up to and including the index date (primary exposure) was compared to non-use, defined as no PPI prescription fill in the 365 days prior to the index date. The primary outcome was a composite of use of mechanical ventilation or death within 60 days. Weighted logistic regression models evaluated severe COVID-19 outcomes between current PPI users vs non-users.

**Results:** Of 97,674 Veterans with SARS-CoV-2 testing, 14,958 tested positive (6262 [41.9%] current PPI users, 8696 [58.1%] non-users) and comprised the analytic cohort. After weighting, all covariates were well-balanced. In the weighted cohort, there was no difference in the primary composite outcome (8.2% vs 8.0%; OR 1.03, 95% CI 0.91-1.16), secondary composite outcome, nor individual component outcomes between current PPI users and non-users. There was no significant interaction between age and PPI use on outcomes.

**Conclusion:** Among patients with SARS-CoV-2 infection, current PPI use vs non-use is not associated with severe COVID-19 outcomes.

## Introduction

Coronavirus disease-2019 (COVID-19), caused by the novel severe acute respiratory syndrome coronavirus-2 (SARS-CoV-2), is responsible for over 3 million deaths worldwide including nearly 600,000 deaths in the United States (US), to date.^1^ Furthermore, while therapies and knowledge of the disease have decreased the mortality rate over time, there remains substantial morbidity associated with COVID-19.^2–4^ However, much remains unknown, and clinical understanding of factors—especially modifiable factors, such as common drug exposures—associated with severe COVID-19 related outcomes is limited.^5–7^

Proton pump inhibitors (PPIs) are among the most commonly prescribed medications in the US and worldwide. PPIs irreversibly bind to the hydrogen-potassium ATPase of parietal cells to suppress gastric acid secretion, causing gastric pH to rise above the standard pH 1.5-3.5.^8^ Most microbes, including coronaviruses, are deactivated at this low pH.^9^ SARS-CoV-2 gains entry into cells via the angiotensin-converting enzyme II (ACE2) receptor, which, in addition to being expressed on type 2 alveolar cells in the lung, is also concentrated on intestinal epithelial cells.^10,11^ Enterocytes are an important site for SARS-CoV-2 replication.^12^ PPI-induced potent gastric acid suppression might allow the ACE2 receptor-expressing enterocytes to be exposed to higher SARS-CoV-2 viral load^9,10^, leading to subsequent downstream consequences that are hypothesized to be related to a larger pathogen burden, such as COVID-19 cytokine storm.^8,13,14^

Given the continued overwhelming burden of COVID-19, the high prevalence of PPI use, especially among US veterans^15^, clarifying the association between PPI use and severe COVID-19 remains a key knowledge gap. The primary objective of this study was to evaluate the hypothesis that current PPI use vs non-use is associated with increased odds of severe COVID-19-related outcomes, defined as the need for mechanical ventilation and death.

## Methods

We conducted a propensity score weighted retrospective analysis using a national veteran cohort. This study was approved by the Veterans Affairs (VA) Tennessee Valley Healthcare system institutional review board and Research and Development Committees with a waiver of informed consent.

### Data Sources and Cohort Development

The Veterans Health Administration (VHA) is the largest integrated health network in the US and provides universal healthcare for more than 9 million US veterans.^16^ The national VHA Observational Medical Outcomes Partnership (OMOP) database contains information from veterans’ electronic health records (EHR), including the Corporate Data Warehouse (CDW), pharmacy files, and the VA Informatics and Computing Infrastructure (VINCI) data warehouse, which aggregates data from all VHA facilities nationwide.^17,18^ From these sources, we extracted Veteran-level data including demographics, prescription details, death dates, and diagnostic and procedure information related to outpatient and inpatient encounters.

The analytic dataset was constructed from a retrospective nationwide cohort of US veterans ≥18 years receiving longitudinal care within the VHA who were tested for *Helicobacter pylori*, which was then linked to the COVID-19 Shared Data Resource (SDR). This population was selected for improved reliability in classifying PPI exposure and to minimize the potential for unmeasured confounding. The COVID-19 SDR is a robust data domain comprising veterans who were tested for SARS-CoV-2. This domain uses case definitions, concept definitions, data mappings, and other information developed from data sources that are verified, validated, and updated collaboratively across the VHA.^17–19^ Data within the COVID-19 SDR continually undergo quality checks for accuracy, with regular data refreshes and manual adjudication. All phenotype algorithms, including for covariates and outcomes, were created using a combination of International Classification of Diseases (ICD) versions 9 (ICD-9) and 10 (ICD-10), Current Procedural Terminology (CPT) codes, natural language processing and keyword text searching of inpatient and outpatient medical encounter notes, pharmacy files, and laboratory tests. Algorithms were validated prior to release in the COVID-19 SDR. Additional details are provided in the **Supplemental Methods**.

### Index date for inclusion in primary analysis, COVID-19-related outcomes

The date of a patient’s first positive test for COVID-19 was the index date (T0) and the start of follow-up (**Figure 1**). Patients without a positive COVID-19 test or available results were excluded. For patients with multiple positive COVID-19 tests, only the first positive result was considered. Only patients with a positive COVID-19 test through January 9, 2021 were included to allow all patients to complete 60 days of follow-up for outcome assessment (study end date was March 10, 2021).

**Figure 1.**
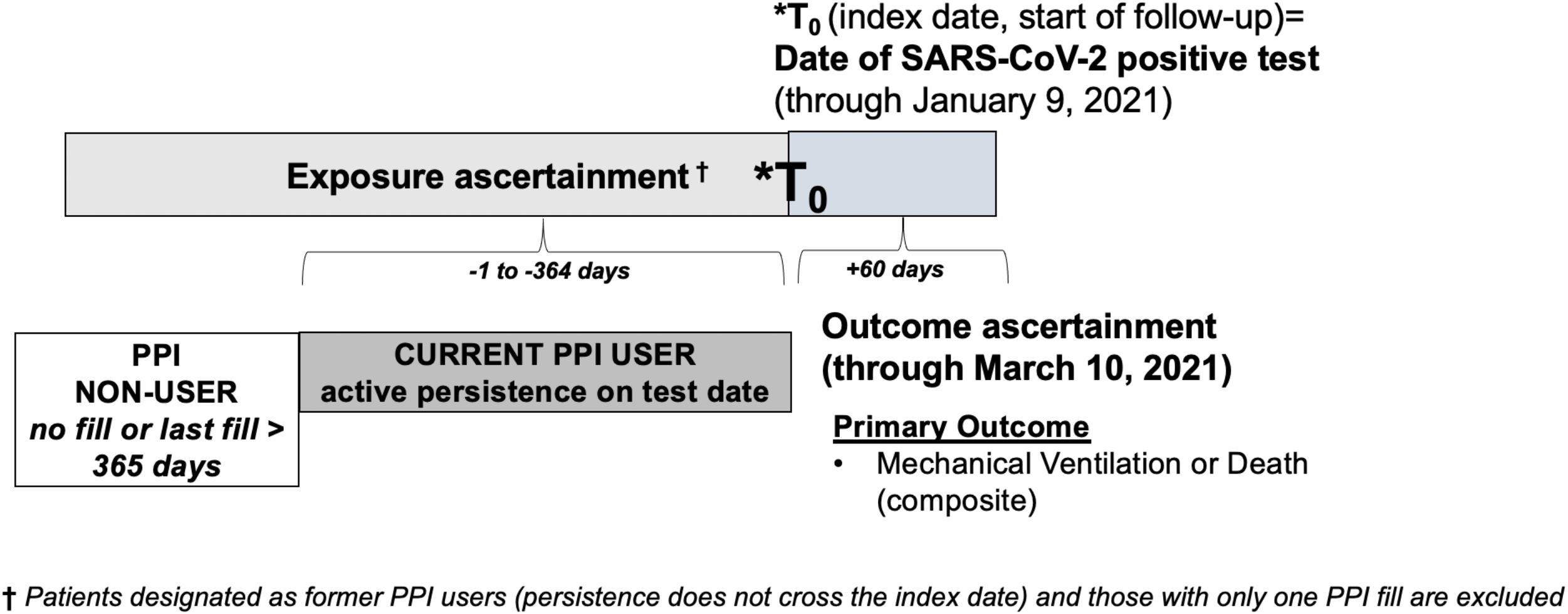
Study design for the primary analysis. The date of a patient’s first positive test for COVID-19 was the index date (T0) and the start of follow-up. Only patients with a positive SARS-CoV-2 test through January 9, 2021 were included to allow all patients to complete 60 days of follow-up for outcome assessment. The primary exposure was current outpatient PPI use up to and including the index date of testing positive for SARS-CoV-2. Patients were categorized as “PPI non-users” if they had not filled an outpatient PPI prescription for at least 365 days prior to the SARS-CoV-2 positive test date. Patients with PPI persistence windows that included -1 to -364 days prior to the index date were considered recent former users and excluded. The primary outcome was a composite of mechanical ventilation or death within 60 days from the index date, which were also analyzed as separate outcomes (see text).

### Exposure: Current PPI use on SARS-CoV-2 test date

The primary exposure was current outpatient PPI use up to and including the index date of testing positive for SARS-CoV-2. Although some PPIs are available over-the-counter, most veterans choose to fill their prescriptions through the VHA given the cost savings.^20,21^ To be eligible for categorization as a current PPI user, a patient needed at least two outpatient PPI prescription fills prior to the index date. The drug persistence was calculated using the dates of the two most recent PPI prescription fills and the dispensed “days supply” of PPI therapy. The days supply was added to the date of the prescription fill, and if this persistence window included the date of the positive SARS-CoV-2 test, the person was categorized as a “current PPI user” (**Figure 1**). Patients were categorized as “PPI non-users” if they had not filled an outpatient PPI prescription for at least 365 days prior to the SARS-CoV-2 positive test date. Patients with PPI persistence windows that included -1 to -364 days prior to the index date were considered recent former users and excluded. To evaluate for protopathic bias among the current PPI users—that is, use of PPIs in response to symptoms that might be the result of COVID-19^22^—we also evaluated the number of days between the first and second PPI prescription fills, as well as the days between the date of the most recent PPI prescription fill and the date of SARS-CoV-2 positive testing.

### Primary and secondary outcomes: COVID-19-related disease severity

The primary outcome was a composite indicator of mechanical ventilation or death within 60 days following the index date. An extended definition included hospitalization and intensive care unit (ICU) admission within 60 days and was the secondary composite outcome. This expanded definition accounts for health system factors, such as bed availability, that may impact hospitalization and ICU admission. Of note, the secondary composite outcome was effectively death and hospitalization since admission to the ICU and mechanical ventilation necessitates hospital admission. All components of composite outcomes were evaluated separately.

### Covariates

Covariates were defined using data available from the two years prior to but not including the index date of SARS-CoV-2 positive testing (−1 to -730 days). We included the following covariates: demographics (VHA facility location, age, sex, and race/ethnicity [categorized as non-Hispanic white, non-Hispanic black, Hispanic, other/unknown]); smoking status (current, former, never, unknown); comorbidities (See **Supplemental Table 1** for definitions of comorbidities); and medications filled in the 90 days prior to the index date (non-steroidal anti-inflammatory drugs (NSAIDs), statins, angiotensin converting enzyme-inhibitors and angiotensin II receptor blockers, histamine-2-receptor antagonists (H2RAs)). To account for temporal trends in COVID-19 epidemiology and treatments, the number of days from January 1, 2020 to the index date was also included as a covariate.

### Statistical Analysis

We compared the means and standard deviations (SD) for continuous variables and proportions for categorical variables between current PPI users and non-users, and characterized these as standardized mean differences (SMDs). SMDs are calculated as the difference between groups in number of standard deviations, and are the preferred measure of covariate balance.^23^ Smaller SMDs indicate better balance between groups.

Propensity score (PS) weighting was used to balance covariate distributions between PPI users and PPI non-users. We calculated the Average Treatment Effect (ATE) weights as the inverse of the probability of the PPI use status observed. Weights were scaled by a constant so that the sum of weights equaled the unweighted cohort’s sample size. To balance the large number of covariates, PS were calculated via cross-validated logistic ridge regression models with optimal shrinkage parameters. To balance nonlinear associations with PPI use, the PS models included restricted cubic splines on all continuous covariates, including the date of positive SARS-CoV-2 testing (index date). By the definitions of the electronic health record-derived comorbidities and cohort entry criteria, no covariates had missing data. Covariate balance in the propensity-weighted cohort was assessed using SMDs, with a threshold of <0.1 indicating adequate balance, and the ATE weights were inspected for outliers. As determined based on sufficient overlap in the PS (**Supplemental Figure 2**), balance was achieved without extreme weights (**Supplemental Figure 3**). In this circumstance, PS weighting, which uses all subjects, is more efficient than one-to-one PS matching and preserves the effective sample size.

Weighted logistic regression models were used for the primary analysis to compare the composite outcome of mechanical ventilation or death within 60 days for current PPI users vs PPI non-users. A similar analytic approach was utilized for all secondary outcomes. We evaluated for interaction by age (age <65 vs ≥65 years at the index date). All analyses were performed in R 4.0.2, and the survey package was used to account for the propensity score weights.^24^

## Results

### Analytic Sample and Characteristics

Among the 97,674 patients in the study cohort with SARS-CoV-2 testing and complete outcomes data, 14,958 patients (15.3%) tested positive for SARS-CoV-2. These predominantly non-Hispanic white male patients had a mean age over 60, 12% were current smokers, and the most common comorbidities included hypertension, diabetes, chronic obstructive lung disease, cerebrovascular disease, and obstructive sleep apnea (**Table 1**). Of these, 6262 [41.9%] were current PPI users and 8696 [58.1%] were PPI non-users (**Figure 2**). Among current PPI users, nearly all (>96%) current users had persistent PPI use with: 2 outpatient PPI prescriptions filled in the 365 days prior to the index date; at least 14 days between their first and second most recent PPI prescriptions; and at least 14 days between their first most recent PPI fill and their index date. The temporal patterns in the rates of SARS-CoV-2 infection among all veterans tested for mirrored national trends^1^, ranging from 6.8% (May, June, September 2020) to 33.7% in December 2020 (January 1-9, 2021, 38.9% for the partial month) (**Supplemental Figure 1**).

**Table 1.** Characteristics of veterans with positive SARS-CoV-2 testing, stratified by current PPI user vs. PPI non-user

| COVARIATES | UNWEIGHTED COHORT of veterans<br>with positive SARS-CoV-2 test |  | WEIGHTED COHORT of veterans with positive<br>SARS-CoV-2 test |  | SMD* |
| --- | --- | --- | --- | --- | --- |
|  | PPI non-users<br>(N=8,696) | Current PPI users<br>(N=6,262) | PPI non-users<br>(N=8,696) | Current PPI users<br>(N=6,262) |  |
| **VHA Facility<sup>^</sup>** | <sup>^</sup> | <sup>^</sup> | <sup>^</sup> | <sup>^</sup> | 0.070 |
| **Age, mean years (SD)** | 60.46 (15.77) | 64.37 (13.42) | 61.94 (15.14) | 62.15 (14.52) | 0.014 |
| **Male sex, n (%)** | 7,382 (84.9) | 5,578 (89.1) | 7,529 (86.6) | 5,441 (86.9) | 0.009 |
| **Race/Ethnicity, n (%)** |  |  |  |  | 0.033 |
| Non-Hispanic White | 4,437 (51.0) | 3,885 (62.0) | 4,757 (54.7) | 3,527 (56.3) |  |
| Non-Hispanic Black | 2,243 (25.8) | 1,315 (21.0) | 2,125 (24.4) | 1,477 (23.6) |  |
| Non-Hispanic other or<br>unknown | 794 (9.1) | 515 (8.2) | 764 (8.8) | 540 (8.6) |  |
| Hispanic | 1,222 (14.1) | 547 (8.7) | 1,049 (12.1) | 717 (11.5) |  |
| **Days from Jan 1, 2020 to**<br>**index date, mean (SD)<sup>#</sup>** | 282 (82.8) | 289 (78.7) | 285 (81.5) | 285 (81.2) | 0.005 |
| **Smoking Status, n (%)** |  |  |  |  | 0.067 |
| Current Smoker | 1,030 (11.8) | 754 (12.0) | 1,047 (12.0) | 762 (12.2) |  |
| Former Smoker | 3,441 (39.6) | 3,085 (49.3) | 3,773 (43.4) | 2,780 (44.4) |  |
| Never Smoker | 3,430 (39.4) | 2,180 (34.8) | 3,260 (37.5) | 2,376 (37.9) |  |
| Unknown | 795 (9.1) | 243 (3.9) | 616 (7.1) | 344 (5.5) |  |
| **Comorbidities, n (%)** |  |  |  |  |  |
| Asthma | 629 (7.2) | 685 (10.9) | 747 (8.6) | 571 (9.1) | 0.018 |
| Coronary Artery Disease | 1,645 (18.9) | 1,911 (30.5) | 2,034 (23.4) | 1,500 (24.0) | 0.013 |
| Cancer | 1,770 (20.4) | 1,750 (27.9) | 2,020 (23.2) | 1,508 (24.1) | 0.020 |
| Cardiomyopathy | 255 (2.9) | 279 (4.5) | 309 (3.5) | 228 (3.6) | 0.005 |
| Charlson Comorbidity Index,<br>mean (SD) | 1.82 (2.22) | 2.55 (2.54) | 2.12 (2.38) | 2.17 (2.40) | 0.022 |
| Congestive Heart Failure | 621 (7.1) | 746 (11.9) | 791 (9.1) | 582 (9.3) | 0.007 |
| Chronic Lung Disease | 2,652 (30.5) | 2,757 (44.0) | 3,129 (36.0) | 2,315 (37.0) | 0.020 |
| Chronic Neuromuscular<br>Disease | 394 (4.5) | 323 (5.2) | 420 (4.8) | 308 (4.9) | 0.004 |
| Chronic Kidney Disease | 1,161 (13.4) | 1,219 (19.5) | 1,390 (16.0) | 1,026 (16.4) | 0.011 |
| Chronic Kidney Failure | 153 (1.8) | 151 (2.4) | 180 (2.1) | 130 (2.1) | 0.001 |
| Chronic Obstructive<br>Pulmonary Disease | 1,316 (15.1) | 1,646 (26.3) | 1,694 (19.5) | 1,270 (20.3) | 0.020 |
| Cerebrovascular Disease | 2,800 (32.2) | 2,966 (47.4) | 3,320 (38.2) | 2,453 (39.2) | 0.021 |
| Diabetes | 3,070 (35.3) | 2,815 (45.0) | 3,415 (39.3) | 2,469 (39.4) | 0.003 |
| Drug Dependency | 379 (4.4) | 345 (5.5) | 416 (4.8) | 311 (5.0) | 0.008 |
| Emphysema | 126 (1.4) | 170 (2.7) | 171 (2.0) | 126 (2.0) | 0.003 |
| Heart Disease | 2,093 (24.1) | 2,281 (36.4) | 2,533 (29.1) | 1,846 (29.5) | 0.008 |
| Heart Failure (non-congestive) | 790 (9.1) | 898 (14.3) | 988 (11.4) | 709 (11.3) | 0.001 |
| *H. pylori* positive | 1,841 (21.2) | 1,138 (18.2) | 1,746 (20.1) | 1,232 (19.7) | 0.022 |
| HIV | 120 (1.4) | 51 (0.8) | 105 (1.2) | 78 (1.2) | 0.003 |
| Hypertension | 5,075 (58.4) | 4,620 (73.8) | 5,616 (64.6) | 4,149 (66.3) | 0.035 |
| Lower Respiratory Infection | 1,010 (11.6) | 855 (13.7) | 1,076 (12.4) | 796 (12.7) | 0.010 |
| Obstructive Sleep Apnea | 2,884 (33.2) | 2,773 (44.3) | 3,254 (37.4) | 2,454 (39.2) | 0.037 |
| **Medications, n (%)** |  |  |  |  |  |
| ACE Inhibitors | 2,233 (25.7) | 2,152 (34.4) | 2,549 (29.3) | 1,887 (30.1) | 0.018 |
| ARBs | 1,170 (13.5) | 1,239 (19.8) | 1,378 (15.8) | 1,036 (16.6) | 0.019 |
| H2RAs | 626 (7.2) | 423 (6.8) | 638 (7.3) | 459 (7.3) | 0.001 |
| NSAIDs | 5,359 (61.6) | 4,745 (75.8) | 5,812 (66.8) | 4,358 (69.6) | 0.059 |
| Statins | 4,176 (48.0) | 4,249 (67.9) | 4,832 (55.6) | 3,656 (58.4) | 0.057 |

Abbreviations: Angiotensin converting enzyme inhibitors (ACE inhibitors); Angiotensin II receptor blockers (ARBs); *Helicobacter pylori* (*H. pylori*); Histamine-2 receptor antagonists (H2RAs); Non-steroidal anti-inflammatory agents (NSAIDs); Proton pump inhibitor (PPI); standard deviation (SD); standardized mean difference (SMD); Veterans Health Administration (VHA)
\*Only SMDs for the weighted cohort are provided in this table. Please refer to Supplemental Figure 4 for the SMD plots for both the unweighted and weighted cohorts.
^All of the 127 VHA facilities were included as covariates in this analysis; however, the proportion of patients at each station for each group is not listed here due to space considerations. The SMD between PPI users and non-users in the unweighted cohort was 0.36, with balance achieved after weighting (SMD 0.07).
#This variable represents the days from January 1, 2020 to the index date of SARS-CoV-2 testing to account for temporal differences. Please refer text for additional details.

**Figure 2.**
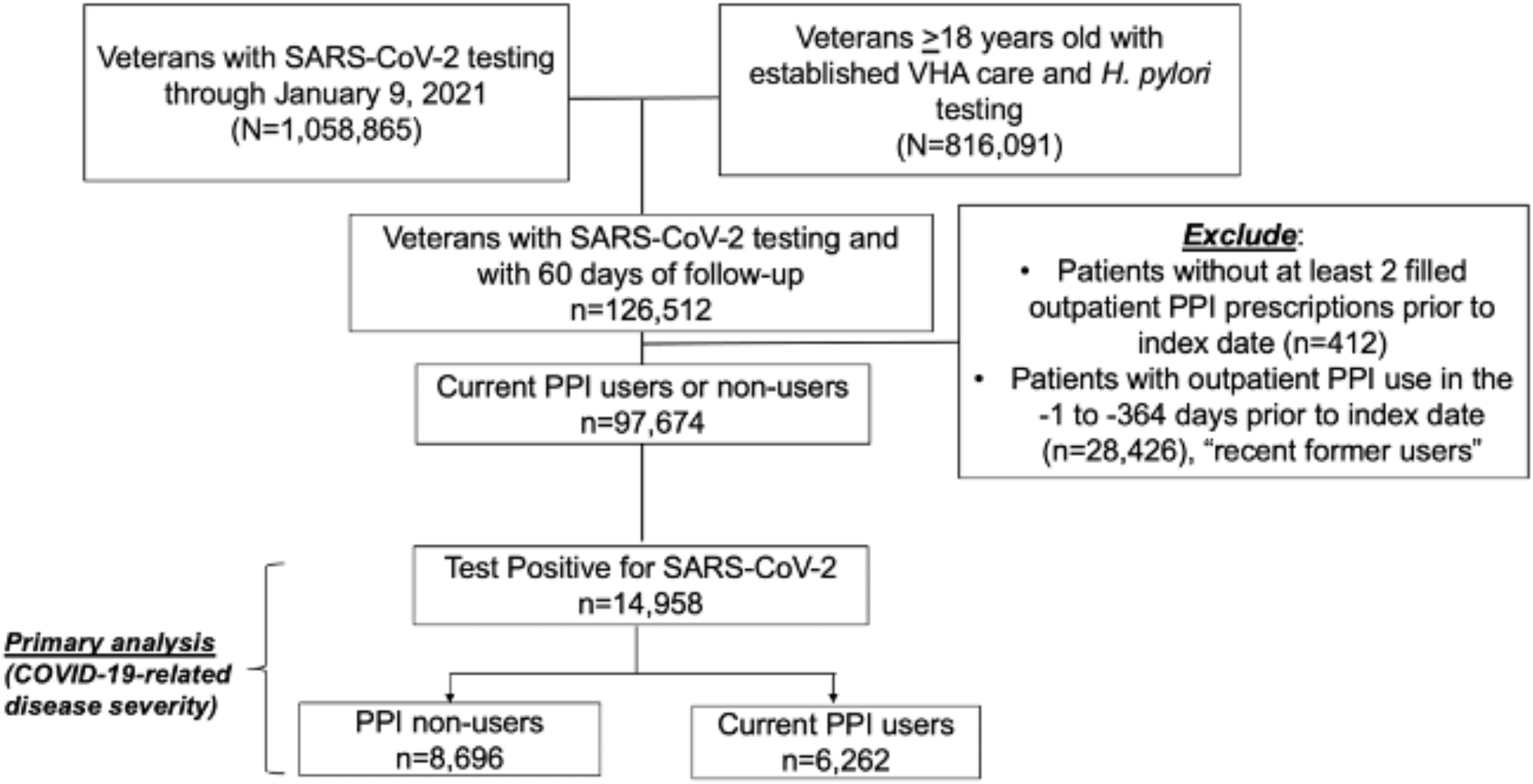
Flow diagram of cohort construction. The analytic dataset was constructed from a retrospective nationwide cohort of US veterans ≥18 years receiving longitudinal care within the VHA who were tested for *Helicobacter pylori*, which was then linked to the COVID-19 Shared Data Resource domain. Only patients with a positive COVID-19 test through January 9, 2021 were included. The primary analytic cohort comprised 14,958 patients who tested positive for SARS-CoV-2, 6,262 of whom were classified as current PPI users and 8,696 as PPI non-users.

In the unweighted cohort, current PPI users were older, more often current or former smokers, and had more comorbidities than PPI non-users. After weighting, all covariates were well-balanced with no SMD greater than 0.06 between the two groups (**Table 1, Supplemental Figure 3**). Patient demographics, comorbidities, and medications stratified by current PPI user vs non-user for the unweighted and weighted cohorts are provided in **Table 1**.

After propensity score weighting, patient-reported COVID-19 symptoms, including GI symptoms, were also similar between current PPI users vs non-users (**Supplemental Table 2** weighted and unweighted cohort).

### Primary analysis: Severe COVID-19 related outcomes among current PPI users vs non-users

In the unweighted cohort of patients with COVID-19, there was a higher frequency and higher odds of the primary composite outcome of mechanical ventilation or death in current PPI users vs non-users (9.3% vs 7.5%; OR 1.27, 95% CI 1.13-1.43) (**Table 2**). The pattern was similar for the secondary composite outcome of hospitalization, ICU admission, mechanical ventilation, or death within 60 days (25.8% vs 21.4%; OR 1.27, 95% CI 1.18-1.37), among current PPI users vs non-users. Individual outcomes were consistent with the composite results (**Table 2, Figure 3**). After propensity score weighting, there was no difference in the primary composite outcome between current PPI users and non-users (composite 8.2% vs 8.0%; OR 1.03, 95% CI 0.91-1.16).

**Table 2.**
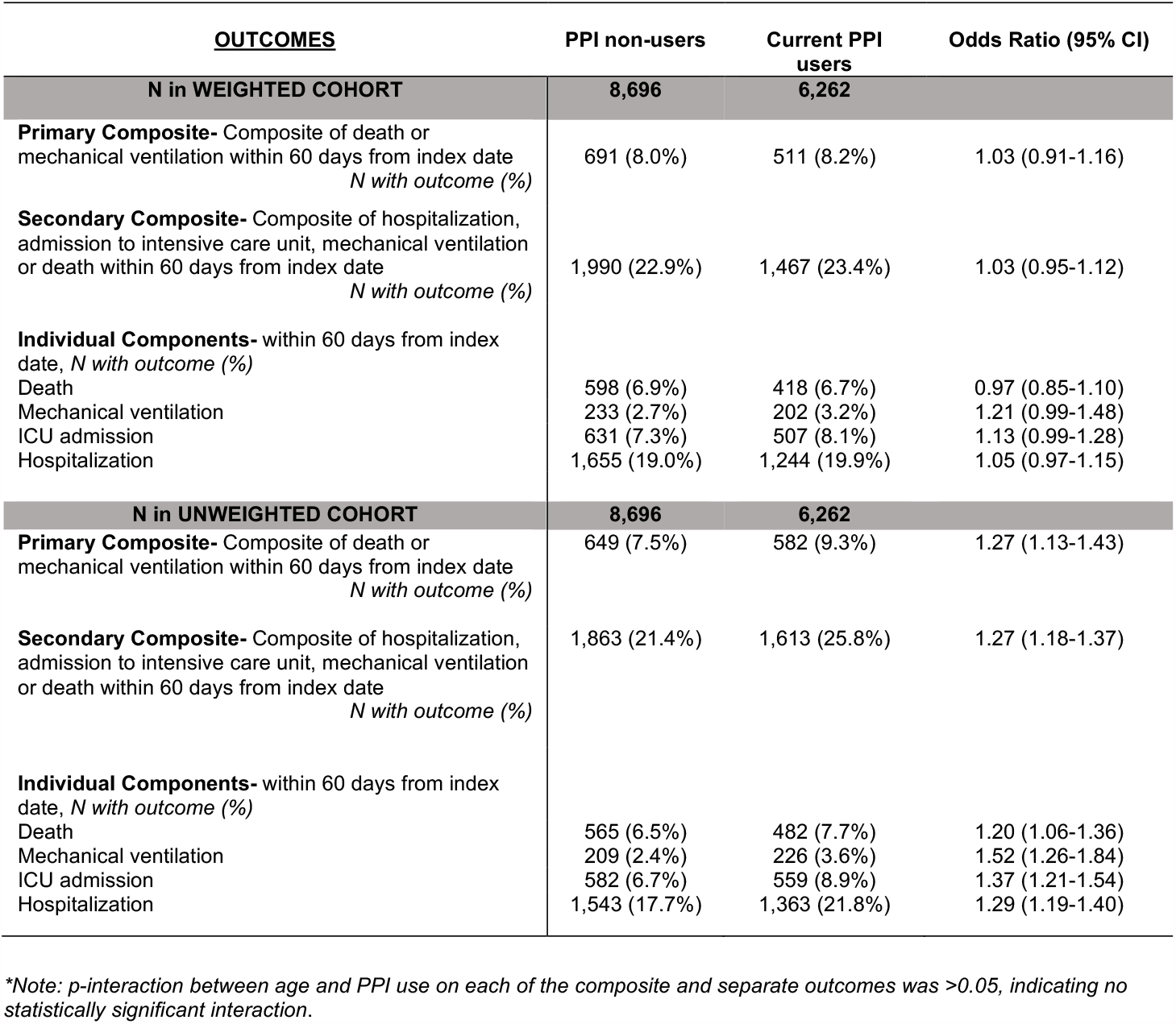
Associations between current PPI use vs. PPI non-use and COVID-19-related disease severity outcomes (primary analysis, unweighted and weighted cohorts)

| **OUTCOMES** | **PPI non-users** | **Current PPI users** | **Odds Ratio (95% CI)** |
| --- | --- | --- | --- |
| **N in WEIGHTED COHORT** | **8,696** | **6,262** |  |
| **Primary Composite-** Composite of death or mechanical ventilation within 60 days from index date<br>*N with outcome (%)* | 691 (8.0%) | 511 (8.2%) | 1.03 (0.91-1.16) |
| **Secondary Composite-** Composite of hospitalization, admission to intensive care unit, mechanical ventilation or death within 60 days from index date<br>*N with outcome (%)* | 1,990 (22.9%) | 1,467 (23.4%) | 1.03 (0.95-1.12) |
| **Individual Components-** within 60 days from index date, *N with outcome (%)* |  |  |  |
| Death | 598 (6.9%) | 418 (6.7%) | 0.97 (0.85-1.10) |
| Mechanical ventilation | 233 (2.7%) | 202 (3.2%) | 1.21 (0.99-1.48) |
| ICU admission | 631 (7.3%) | 507 (8.1%) | 1.13 (0.99-1.28) |
| Hospitalization | 1,655 (19.0%) | 1,244 (19.9%) | 1.05 (0.97-1.15) |
| **N in UNWEIGHTED COHORT** | **8,696** | **6,262** |  |
| **Primary Composite-** Composite of death or mechanical ventilation within 60 days from index date<br>*N with outcome (%)* | 649 (7.5%) | 582 (9.3%) | 1.27 (1.13-1.43) |
| **Secondary Composite-** Composite of hospitalization, admission to intensive care unit, mechanical ventilation or death within 60 days from index date<br>*N with outcome (%)* | 1,863 (21.4%) | 1,613 (25.8%) | 1.27 (1.18-1.37) |
| **Individual Components-** within 60 days from index date, *N with outcome (%)* |  |  |  |
| Death | 565 (6.5%) | 482 (7.7%) | 1.20 (1.06-1.36) |
| Mechanical ventilation | 209 (2.4%) | 226 (3.6%) | 1.52 (1.26-1.84) |
| ICU admission | 582 (6.7%) | 559 (8.9%) | 1.37 (1.21-1.54) |
| Hospitalization | 1,543 (17.7%) | 1,363 (21.8%) | 1.29 (1.19-1.40) |

\*Note: *p*-interaction between age and PPI use on each of the composite and separate outcomes was >0.05, indicating no statistically significant interaction.

**Figure 3.**
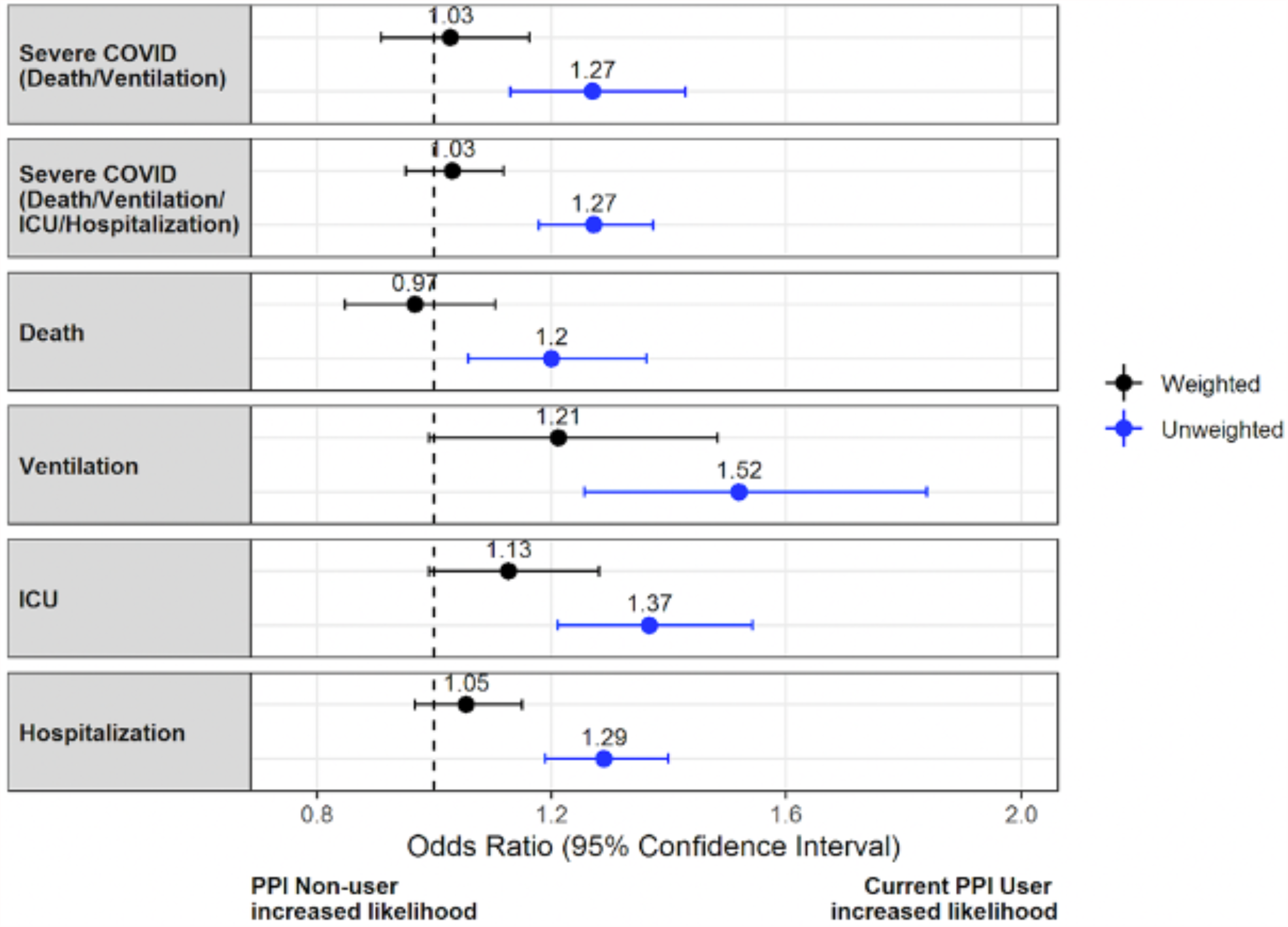
Forest plot of primary and secondary COVID-19 outcomes within 60 days of the index date, weighted and unweighted cohorts. In the unweighted cohort, current outpatient PPI use compared to PPI non-use was associated with increased odds of severe COVID-19 outcomes, defined based on composite (primary: death or mechanical ventilation; secondary: death, mechanical ventilation, intensive care unit admission, or hospitalization) and individual component outcomes. Each of these associations were statistically nonsignificant after more fully accounting for covariates in the propensity-weighted cohort, including date of SARS-CoV-2 testing and VHA facility location. Of note, there was no significant interaction between age group and PPI use on these outcomes (see text).

Results were consistent for the secondary composite outcome in the weighted analysis (23.4% vs 22.9%, OR 1.03, 95% CI 0.95-1.12), and individual component outcomes (**Table 2, Figure 3**). Of note, the frequency of dexamethasone use for COVID-19 treatment was similar between current PPI users and PPI non-users in the weighted cohort (12.7% vs 11.5%).

There was no statistically significant interaction between age and PPI use on COVID-19 severity outcomes (all interaction P values >0.05).

## Discussion

We demonstrated that there are no increased odds of severe COVID-19 outcomes associated with current outpatient PPI use compared to non-use in this large nationwide propensity-weighted cohort analysis. In an unweighted analysis, we observed a significantly increased odds of severe outcomes with PPI use, similar to previously published studies that reported minimally adjusted results. However, these associations became statistically nonsignificant after more fully accounting for covariates in the propensity-weighted cohort, including date of SARS-CoV-2 testing as well as VHA facility location. No significant interaction between age group and PPI use on these outcomes was observed in composites or individually.

To date, several studies analyzing COVID-19-related outcomes in patients categorized as PPI users vs non-users demonstrate mixed results.^25–28^ These varied findings in large part reflect heterogeneity across studies with respect to definitions of PPI exposure and COVID-19 severity outcomes, rigor in exposure and outcome assessment, covariate assessment and adjustment, study design, sample size, and heterogeneity across study populations with respect to geography, demographic distributions, COVID-19 prevalence, contemporaneous treatments and healthcare infrastructure. All prior studies included data from the first few months of the global COVID-19 global pandemic, which represent the months when our understanding of COVID-19 management and available treatments was rapidly evolving. This study represents more than a full year of the pandemic, is national in scope, standardized the primary outcome definition and accounted for comorbidities, as well as the dynamic pandemic timeframe with respect to COVID-19 epidemiology, geographic changes and clinical management evolution.

At least four separate meta-analyses demonstrated significantly increased risk of severe COVID-19-related outcomes among PPI users compared to non-users,^25–28^ typically defined as ICU admission, mechanical ventilation or death. The meta-analyses largely included studies of small sample size which did not adjust for important comorbidities, such as tobacco use. In the unweighted cohort for the present analysis, we also observed an association between PPI use and severe COVID-19 outcomes (death, mechanical ventilation, ICU admission, and hospitalization separately and as composites) which was not demonstrated in the propensity score-weighted cohort, suggesting that the associations in previous studies reflect incomplete covariate adjustment.^22^ Notably, while an association between PPI use vs non-use and severe COVID-19 outcomes was demonstrated in the REACT-SCOT case-control study, this association was attenuated when considering PPI exposure in a 120-day window prior to COVID-19 diagnosis.^29^ Strict definitions of medication exposure windows are needed to accurately assess safety outcomes.^30^ In the present study, we excluded patients who: 1) did not have at least 2 filled outpatient PPI prescriptions prior to the index date; and 2) were categorized as recent former users (i.e. their PPI persistence window did not overlap with the index date). The strict exposure ascertainment window reduces misclassification. Nearly all individuals classified as current PPI users in our cohort filled ≥2 PPI prescriptions within 1 year from the index date and had >14 days between their 2 most recent PPI fills and also between their most recent PPI fill and index date, indicating persistent PPI use and also minimizing the likelihood of protopathic bias. That self-reported GI symptoms in the 30 days prior to SARS-CoV-2 testing were similar between current PPI users and non-users also minimizes protopathic bias.

The study strengths include its national scope, large size and year-long duration. We utilized robust, verified national VHA data and achieved complete covariate balance to minimize possible confounding due to comorbidities, smoking, concomitant medication use including H2RAs, VHA facility, and date of testing. All patients included were established in VHA care and had complete data for covariates. We reproduced the findings demonstrated in prior studies among the patients in the unweighted cohort, underscoring the impact of potential confounding. With increased underlying comorbidities and lifestyle factors relative to the US population, veterans represent an enriched at-risk population; the absence of increased odds of severe COVID-19 outcomes associated with PPI use is therefore particularly reassuring. Our retrospective study is limited by the possibility for residual confounding. Our findings may not be generalizable to the broader population. Notably, while the overall VHA population is predominantly older white men, the cohort for the primary analysis was comprised of nearly 25% blacks and 10% Hispanics, which reflects the racial and ethnic disparities that underlie the COVID-19 pandemic. As some PPIs are available over-the-counter, the study is limited by the possibility for misclassification of current PPI users as non-users. However, this misclassification may be less likely in the VHA population since veterans can fill medications for free or for very low cost through the VHA.^20,21^ Non-VHA medical care is not captured in the national VHA database. It is therefore possible that veterans who tested positive for SARS-CoV-2 and were hospitalized outside of VHA are missed in this analysis. Notably, age, which is the primary driver of non-VHA care due to Medicare eligibility, was balanced between current PPI users and non-users. Therefore, we have no strong reason to believe that outcomes occurred differentially. Moreover, there was no significant interaction between age group and PPI use on any of the COVID-19-related individual and composite outcomes.

In conclusion, this national comprehensive propensity score-weighted VHA analysis demonstrated no association between current PPI use and severe COVID-19 outcomes. With respect to COVID-19, patients and providers should feel safe to continue to use PPIs at the lowest effective dose for approved indications.

## Data Availability

Data used for this analysis reside in the Veterans Health Administration database. Only approved VA investigators can access the data in order to inform their currently approved study proposals. The data undergo routine quality checks. These data are not and cannot be made publicly available.

## Acknowledgement Section

This research is based on data from the Veterans Health Administration including the VA COVID-19 Shared Data Resource Domain and supported by the Office of Research and Development. This publication does not represent the views of the Department of Veteran Affairs or the United States Government. We also acknowledge the support of the members of the Million Veteran Program Pharmacogenomics Working Group.

## Author contributions

SCS: Study concept, study design, dataset verification, interpretation of data and statistical analysis, drafting of initial manuscript, critical revision of manuscript; AEH, RAG: study design, primary statistical analysis, dataset verification, interpretation of data and statistical analysis, critical revision of manuscript, methodological oversight; CD, OW, JD: dataset creation and stewardship; BM, ST, KC, AS, CMH, ES, MEM, AH: manuscript revision; CLR: study design, interpretation of data and statistical analysis, methodological and study oversight, critical revision of manuscript. All authors approved the final version of the manuscript.

## Figure Legends

**Supplemental Figure 1. Proportion of SARS-CoV-2 positive testing among veterans tested through January 9, 2021**. Histogram illustrating the number of SARS-CoV-2 tests performed within the VHA nationwide between March 1, 2020 through January 9, 2020, and the proportion of positive test results. The magnitude and temporality of the SARS-CoV-2 positivity rate reflects the overall observed trend nationally during this time period.

**Supplemental Figure 2. Distribution of propensity scores for current PPI users vs. non-users among veterans who tested positive for SARS-CoV-2 (N=14**,**958**; **primary analytic cohort)**. Propensity score (PS) weighting was used to balance covariate distributions between PPI users and PPI non-users. We calculated the Average Treatment Effect (ATE) weights as the inverse of the probability of the observed PPI use statuses. Weights were scaled by a constant so that the sum of weights equaled the unweighted cohort’s sample size. To balance the large number of covariates, PS were calculated via cross-validated logistic ridge regression models with optimal shrinkage parameters. To balance nonlinear associations with PPI use, the PS models included restricted cubic splines on all continuous covariates, including the date of positive SARS-CoV-2 testing (index date). As determined based on sufficient overlap in the PS, balance was achieved without extreme weights (see Supplemental Figure 3)

**Supplemental Figure 3. Plot of standardized mean differences for covariates between current PPI users and non-users for the unweighted and weighted cohorts**. Standardized mean differences (SMDs) were used to compare the means and standard deviations (SD) for continuous variables and proportions for categorical variables between current PPI users and non-users. SMDs are the preferred measure of covariate balance in large cohorts. Smaller SMDs indicate better balance between groups, with a threshold of <0.1 (red vertical line) indicating adequate balance. As depicted in the SMD plot of both the unweighted and propensity score-weighted cohort, full covariate balance was achieved in the weighted cohort.

