## Supplemental Material for "Proton pump inhibitor use is not associated with severe COVID-19 related outcomes: A propensity score weighted analysis of a national veteran cohort"

**Supplemental Material – Table of Contents**

Page 1 – Supplemental Methods
Page 2 – Supplemental Table 1
Page 14 – Supplemental Table 2
Page 15 – Supplemental Table 3
Page 16 – Supplemental Figure 1
Page 17 – Supplemental Figure 2
Page 18 – Supplemental Figure 3

**Supplemental Methods,** *Additional details regarding the VA COVID-19 Shared Data Resource Domain*

The COVID-19 SDR is a robust data domain that uses case definitions, concept definitions, data mappings, and other information developed from various data sources, all of which are verified, validated, and updated collaboratively across the VHA. The COVID-19 SDR domain includes the following data specifically related to COVID-19, which are defined using a combination of International Classification of Diseases (ICD) versions 9 (ICD-9) and 10 (ICD-10) and Current Procedural Terminology (CPT) codes, natural language processing/keyword text searching of inpatient and outpatient medical encounter notes, pharmacy files, and laboratory tests: confirmed SARS-CoV-2 testing (based on laboratory tests that comply with the Centers for Disease Control and Prevention [CDC] standards, such as the 2019-nCOV RT-PCR Diagnostic Panel and the SARS-CoV-2 Multiplex Assay, or human case review) and results, detailed demographics, extensive covariates, such as comorbidities and prior/current medications, COVID-19 related symptoms (asymptomatic, loss of smell or taste, general systemic, respiratory, or gastrointestinal symptoms), as well as treatments and health-related outcomes, among other Veteran-level details. The VHA's National Surveillance Tool provides near real-time information on all SARS-CoV-2 cases including hospitalization course and outcomes, ensuring that all data are up to date. Data contained within the COVID-19 SDR continually undergo quality checks for accuracy and data refreshes. All phenotype algorithms were validated prior to release in the COVID SDR and are vetted by human review. The data for this study were up to date through March 10, 2021.

**Supplemental Table 1. Covariate definitions***

| | Definition |
|---|---|
| **Conditions** | |
| Asthma | ICD9CM: 493, 493.01, 493.02, 493.1, 493.11, 493.12, 493.2, 493.21, 493.22, 493.81, 493.82, 493.9, 493.91, 493.92<br><br>ICD10CM: J45.20, J45.21, J45.22, J45.30, J45.31, J45.32, J45.40, J45.41, J45.42, J45.50, J45.51, J45.52, J45.901, J45.902, J45.909, J45.991, J45.998, J82 |
| Cancer | ICD9CM: 84, 84.2, 84.3, 84.4, 84.5, 84.6, 84.7, 84.8, 84.9, 140, 140.1, 140.3, 140.4, 140.5, 140.6, 140.8, 140.9, 141, 141.1, 141.2, 141.3, 141.4, 141.5, 141.6, 141.8, 141.9, 142, 142.1, 142.2, 142.8, 142.9, 143, 143.1, 143.8, 143.9, 144, 144.1, 144.8, 144.9, 145, 145.1, 145.2, 145.3, 145.4, 145.5, 145.6, 145.8, 145.9, 146, 146.1, 146.2, 146.3, 146.4, 146.5, 146.6, 146.7, 146.8, 146.9, 147, 147.1, 147.2, 147.3, 147.8, 147.9, 148, 148.1, 148.2, 148.3, 148.8, 148.9, 149, 149.1, 149.8, 149.9, 150, 150.1, 150.2, 150.3, 150.4, 150.5, 150.8, 150.9, 151, 151.1, 151.2, 151.3, 151.4, 151.5, 151.6, 151.8, 151.9, 152, 152.1, 152.2, 152.3, 152.8, 152.9, 153, 153.1, 153.2, 153.3, 153.4, 153.5, 153.6, 153.7, 153.8, 153.9, 154, 154.1, 154.2, 154.3, 154.8, 155, 155.1, 155.2, 156, 156.1, 156.2, 156.8, 156.9, 157, 157.1, 157.2, 157.3, 157.4, 157.8, 157.9, 158, 158.8, 158.9, 159, 159.1, 159.8, 159.9, 160, 160.1, 160.2, 160.3, 160.4, 160.5, 160.8, 160.9, 161, 161.1, 161.2, 161.3, 161.8, 161.9, 162, 162.2, 162.3, 162.4, 162.5, 162.8, 162.9, 163, 163.1, 163.8, 163.9, 164, 164.1, 164.2, 164.3, 164.8, 164.9, 165, 165.8, 165.9, 170, 170.1, 170.2, 170.3, 170.4, 170.5, 170.6, 170.7, 170.8, 170.9, 171, 171.2, 171.3, 171.4, 171.5, 171.6, 171.7, 171.8, 171.9, 173.1, 173.11, 173.12, 173.19, 174, 174.1, 174.2, 174.3, 174.4, 174.5, 174.6, 174.8, 174.9, 175, 175.9, 179, 180, 180.1, 180.8, 180.9, 181, 182, 182.1, 182.8, 183, 183.2, 183.3, 183.4, 183.5, 183.8, 183.9, 184, 184.1, 184.2, 184.3, 184.4, 184.8, 184.9, 185, 186, 186.9, 187.1, 187.2, 187.3, 187.4, 187.5, 187.6, 187.7, 187.8, 187.9, 188, 188.1, 188.2, 188.3, 188.4, 188.5, 188.6, 188.7, 188.8, 188.9, 189.1, 189.2, 189.3, 189.4, 189.8, 189.9, 190, 190.1, 190.2, 190.3, 190.4, 190.5, 190.6, 190.7, 190.8, 190.9, 191, 191.1, 191.2, 191.3, 191.4, 191.5, 191.6, 191.7, 191.8, 191.9, 192, 192.1, 192.2, 192.3, 192.8, 192.9, 193, 194, 194.1, 194.3, 194.4, 194.5, 194.6, 194.8, 194.9, 195, 195.1, 195.2, 195.3, 195.4, 195.5, 195.8, 196, 196.1, 196.2, 196.3, 196.5, 196.6, 196.8, 196.9, 197, 197.1, 197.2, 197.3, 197.4, 197.5, 197.6, 197.7, 197.8, 198.1, 198.3, 198.4, 198.5, 198.6, 198.7, 198.81, 198.82, 198.89, 199, 199.1, 199.2, 200.2, 200.21, 200.22, 200.23, 200.24, 200.25, 200.26, 200.27, 200.28, 200.3, 200.31, 200.32, 200.33, 200.34, 200.35, 200.36, 200.37, 200.38, 200.4, 200.41, 200.42, 200.43, 200.44, 200.45, 200.46, 200.47, 200.48, 200.5, 200.51, 200.52, 200.53, 200.54, 200.55, 200.56, 200.57, 200.58, 200.6, 200.61, 200.62, 200.63, 200.64, 200.65, 200.66, 200.67, 200.68, 200.7, 200.71, 200.72, 200.73, 200.74, 200.75, 200.76, 200.77, 200.78, 201.4, 201.41, 201.42, 201.43, 201.44, 201.45, 201.46, 201.47, 201.48, 201.5, 201.51, 201.52, 201.53, 201.54, 201.55, 201.56, 201.57, 201.58, 201.6, 201.61, 201.62, 201.63, 201.64, 201.65, 201.66, 201.67, 201.68, 201.7, 201.71, 201.72, 201.73, 201.74, 201.75, 201.76, 201.77, 201.78, 202, 202.01, |

| | |
|---|---|
| | 202.02, 202.03, 202.04, 202.05, 202.06, 202.07, 202.08, 202.1, 202.11, 202.12, 202.13, 202.14, 202.15, 202.16, 202.17, 202.18, 202.2, 202.21, 202.22, 202.23, 202.24, 202.25, 202.26, 202.27, 202.28, 202.3, 202.31, 202.32, 202.33, 202.34, 202.35, 202.36, 202.37, 202.38, 202.4, 202.41, 202.42, 202.43, 202.44, 202.45, 202.46, 202.47, 202.48, 202.5, 202.51, 202.52, 202.53, 202.54, 202.55, 202.56, 202.57, 202.58, 202.6, 202.61, 202.62, 202.63, 202.64, 202.65, 202.66, 202.67, 202.68, 202.7, 202.71, 202.72, 202.73, 202.74, 202.75, 202.76, 202.77, 202.78, 202.8, 202.81, 202.82, 202.83, 202.84, 202.85, 202.86, 202.87, 202.88, 202.9, 202.91, 202.92, 202.93, 202.94, 202.95, 202.96, 202.97, 202.98, 203.1, 203.11, 203.12, 204, 204, 204.01, 204.02, 204.1, 204.1, 204.11, 204.12, 204.2, 204.2, 204.21, 204.22, 204.8, 204.8, 204.81, 204.82, 204.9, 204.9, 204.91, 204.92, 205, 205, 205.01, 205.02, 205.1, 205.1, 205.11, 205.12, 205.2, 205.2, 205.21, 205.22, 205.3, 205.3, 205.31, 205.32, 205.8, 205.8, 205.81, 205.82, 205.9, 205.9, 205.91, 205.92, 206, 206, 206.01, 206.02, 206.1, 206.1, 206.11, 206.12, 206.2, 206.2, 206.21, 206.22, 206.8, 206.8, 206.81, 206.82, 206.9, 206.9, 206.91, 206.92, 207, 207, 207.01, 207.02, 207.1, 207.1, 207.11, 207.12, 207.2, 207.2, 207.21, 207.22, 207.8, 207.8, 207.81, 207.82, 208, 208, 208.01, 208.02, 208.1, 208.1, 208.11, 208.12, 208.2, 208.2, 208.21, 208.22, 208.8, 208.8, 208.81, 208.82, 208.9, 208.9, 208.91, 208.92, 209, 209.01, 209.02, 209.03, 209.1, 209.11, 209.12, 209.13, 209.14, 209.15, 209.16, 209.17, 209.2, 209.21, 209.22, 209.23, 209.24, 209.25, 209.26, 209.27, 209.29, 209.3, 209.31, 209.32, 209.33, 209.34, 209.35, 209.36, 209.7, 209.71, 209.72, 209.73, 209.74, 209.75, 209.79, 230, 230.1, 230.2, 230.3, 230.4, 230.5, 230.6, 230.7, 230.8, 230.9, 231, 231.1, 231.2, 231.8, 231.9, 232.1, 233, 233.1, 233.2, 233.3, 233.3, 233.31, 233.32, 233.39, 233.4, 233.5, 233.6, 233.7, 233.9, 234, 234.8, 234.9, 235, 235.1, 235.2, 235.3, 235.4, 235.5, 235.6, 235.7, 235.8, 235.9, 236, 236.1, 236.2, 236.3, 236.4, 236.5, 236.6, 236.7, 236.9, 236.91, 236.99, 237, 237.1, 237.2, 237.3, 237.4, 237.5, 237.6, 237.7, 237.7, 237.71, 237.72, 237.73, 237.79, 237.9, 238, 238.1, 238.3, 238.4, 238.5, 238.6, 238.7, 238.71, 238.72, 238.73, 238.74, 238.75, 238.76, 238.77, 238.79, 238.8, 238.9, 333.92, 357.3, 380.14, 402, 402.01, 402.1, 402.11, 402.9, 402.91, 403.01, 404.01, 404.02, 404.03, 405.01, 405.09, 511.81, 789.51, 995.86, V84.01, V84.02, V84.03, V84.04, V84.09<br><br>ICD10CM: C00.0, C00.1, C00.2, C00.3, C00.4, C00.5, C00.6, C00.8, C00.9, C01., C02.0, C02.1, C02.2, C02.3, C02.4, C02.8, C02.9, C03.0, C03.1, C03.9, C04.0, C04.1, C04.8, C04.9, C05.0, C05.1, C05.2, C05.8, C05.9, C06.0, C06.1, C06.2, C06.80, C06.89, C06.9, C07., C08.0, C08.1, C08.9, C09.0, C09.1, C09.8, C09.9, C10.0, C10.1, C10.2, C10.3, C10.4, C10.8, C10.9, C11.0, C11.1, C11.2, C11.3, C11.8, C11.9, C12., C13.0, C13.1, C13.2, C13.8, C13.9, C14.0, C14.2, C14.8, C15.3, C15.4, C15.5, C15.8, C15.9, C16.0, C16.1, C16.2, C16.3, C16.4, C16.5, C16.6, C16.8, C16.9, C17.0, C17.1, C17.2, C17.3, C17.8, C17.9, C18.0, C18.1, C18.2, C18.3, C18.4, C18.5, C18.6, C18.7, C18.8, C18.9, C19., C20., C21.0, C21.1, C21.2, C21.8, C22.0, C22.1, C22.2, C22.3, C22.4, C22.7, C22.8, C22.9, C23., C24.0, C24.1, C24.8, C24.9, C25.0, C25.1, C25.2, C25.3, C25.4, C25.7, C25.8, C25.9, C26.0, |

| | |
|---|---|
| | C26.1, C26.9, C30.0, C30.1, C31.0, C31.1, C31.2, C31.3, C31.8, C31.9, C32.0, C32.1, C32.2, C32.3, C32.8, C32.9, C33., C34.00, C34.01, C34.02, C34.10, C34.11, C34.12, C34.2, C34.30, C34.31, C34.32, C34.80, C34.81, C34.82, C34.90, C34.91, C34.92, C37., C38.0, C38.1, C38.2, C38.3, C38.4, C38.8, C39.0, C39.9, C40.00, C40.01, C40.02, C40.10, C40.11, C40.12, C40.20, C40.21, C40.22, C40.30, C40.31, C40.32, C40.80, C40.81, C40.82, C40.90, C40.91, C40.92, C41.0, C41.1, C41.2, C41.3, C41.4, C41.9, C43.0, C43.10, C43.11, C43.111, C43.112, C43.12, C43.121, C43.122, C43.20, C43.21, C43.22, C43.30, C43.31, C43.39, C43.4, C43.59, C43.60, C43.61, C43.62, C43.70, C43.71, C43.72, C45.0, C45.1, C45.2, C45.7, C45.9, C46.1, C46.2, C46.3, C46.4, C46.50, C46.51, C46.52, C46.7, C46.9, C47.0, C47.10, C47.11, C47.12, C47.20, C47.21, C47.22, C47.3, C47.4, C47.5, C47.6, C47.8, C47.9, C48.0, C48.1, C48.2, C48.8, C49.0, C49.10, C49.11, C49.12, C49.20, C49.21, C49.22, C49.3, C49.4, C49.5, C49.6, C49.8, C49.9, C49.A0, C49.A1, C49.A2, C49.A3, C49.A4, C49.A5, C49.A9, C4A.0, C4A.10, C4A.11, C4A.111, C4A.112, C4A.12, C4A.121, C4A.122, C4A.20, C4A.21, C4A.22, C4A.30, C4A.31, C4A.39, C4A.4, C4A.59, C4A.60, C4A.61, C4A.62, C4A.70, C4A.71, C4A.72, C4A.8, C4A.9, C50.011, C50.012, C50.019, C50.021, C50.022, C50.029, C50.111, C50.112, C50.119, C50.121, C50.122, C50.129, C50.211, C50.212, C50.219, C50.221, C50.222, C50.229, C50.311, C50.312, C50.319, C50.321, C50.322, C50.329, C50.411, C50.412, C50.419, C50.421, C50.422, C50.429, C50.511, C50.512, C50.519, C50.521, C50.522, C50.529, C50.611, C50.612, C50.619, C50.621, C50.622, C50.629, C50.811, C50.812, C50.819, C50.821, C50.822, C50.829, C50.911, C50.912, C50.919, C50.921, C50.922, C50.929, C51.0, C51.1, C51.2, C51.8, C51.9, C52., C53.0, C53.1, C53.8, C53.9, C54.0, C54.1, C54.2, C54.3, C54.8, C54.9, C55., C56.1, C56.2, C56.9, C57.00, C57.01, C57.02, C57.10, C57.11, C57.12, C57.20, C57.21, C57.22, C57.3, C57.4, C57.7, C57.8, C57.9, C58., C60.0, C60.1, C60.2, C60.8, C60.9, C61., C62.00, C62.01, C62.02, C62.10, C62.11, C62.12, C62.90, C62.91, C62.92, C63.00, C63.01, C63.02, C63.10, C63.11, C63.12, C63.2, C63.7, C63.8, C63.9, C64.1, C64.2, C64.9, C65.1, C65.2, C65.9, C66.1, C66.2, C66.9, C67.0, C67.1, C67.2, C67.3, C67.4, C67.5, C67.6, C67.7, C67.8, C67.9, C68.0, C68.1, C68.8, C68.9, C69.00, C69.01, C69.02, C69.10, C69.11, C69.12, C69.20, C69.21, C69.22, C69.30, C69.31, C69.32, C69.40, C69.41, C69.42, C69.50, C69.51, C69.52, C69.60, C69.61, C69.62, C69.80, C69.81, C69.82, C69.90, C69.91, C69.92, C70.0, C70.1, C70.9, C71.0, C71.1, C71.2, C71.3, C71.4, C71.5, C71.6, C71.7, C71.8, C71.9, C72.0, C72.1, C72.20, C72.21, C72.22, C72.30, C72.31, C72.32, C72.40, C72.41, C72.42, C72.50, C72.59, C72.9, C73., C74.00, C74.01, C74.02, C74.10, C74.11, C74.12, C74.90, C74.91, C74.92, C75.0, C75.1, C75.2, C75.3, C75.4, C75.5, C75.8, C75.9, C76.0, C76.1, C76.2, C76.3, C76.40, C76.41, C76.42, C76.50, C76.51, C76.52, C76.8, C77.0, C77.1, C77.2, C77.3, C77.4, C77.5, C77.8, C77.9, C78.00, C78.01, C78.02, C78.1, C78.2, C78.30, C78.39, C78.4, C78.5, C78.6, C78.7, C78.80, C78.89, C79.00, C79.01, C79.02, C79.10, C79.11, C79.19, C79.31, C79.32, C79.40, C79.49, C79.51, |

| | |
|---|---|
| | C79.52, C79.60, C79.61, C79.62, C79.70, C79.71, C79.72, C79.81, C79.82, C79.89, C79.9, C7A.00, C7A.010, C7A.011, C7A.012, C7A.019, C7A.020, C7A.021, C7A.022, C7A.023, C7A.024, C7A.025, C7A.026, C7A.029, C7A.090, C7A.091, C7A.092, C7A.093, C7A.094, C7A.095, C7A.096, C7A.098, C7A.1, C7A.8, C7B.00, C7B.01, C7B.02, C7B.03, C7B.04, C7B.09, C7B.1, C7B.8, C80.0, C80.1, C80.2, C81.00, C81.01, C81.02, C81.03, C81.04, C81.05, C81.06, C81.07, C81.08, C81.09, C81.10, C81.11, C81.12, C81.13, C81.14, C81.15, C81.16, C81.17, C81.18, C81.19, C81.20, C81.21, C81.22, C81.23, C81.24, C81.25, C81.26, C81.27, C81.28, C81.29, C81.30, C81.31, C81.32, C81.33, C81.34, C81.35, C81.36, C81.37, C81.38, C81.39, C81.40, C81.41, C81.42, C81.43, C81.44, C81.45, C81.46, C81.47, C81.48, C81.49, C81.70, C81.71, C81.72, C81.73, C81.74, C81.75, C81.76, C81.77, C81.78, C81.79, C81.90, C81.91, C81.92, C81.93, C81.94, C81.95, C81.96, C81.97, C81.98, C81.99, C82.00, C82.01, C82.02, C82.03, C82.04, C82.05, C82.06, C82.07, C82.08, C82.09, C82.10, C82.11, C82.12, C82.13, C82.14, C82.15, C82.16, C82.17, C82.18, C82.19, C82.20, C82.21, C82.22, C82.23, C82.24, C82.25, C82.26, C82.27, C82.28, C82.29, C82.30, C82.31, C82.32, C82.33, C82.34, C82.35, C82.36, C82.37, C82.38, C82.39, C82.40, C82.41, C82.42, C82.43, C82.44, C82.45, C82.46, C82.47, C82.48, C82.49, C82.50, C82.51, C82.52, C82.53, C82.54, C82.55, C82.56, C82.57, C82.58, C82.59, C82.60, C82.61, C82.62, C82.63, C82.64, C82.65, C82.66, C82.67, C82.68, C82.69, C82.80, C82.81, C82.82, C82.83, C82.84, C82.85, C82.86, C82.87, C82.88, C82.89, C82.90, C82.91, C82.92, C82.93, C82.94, C82.95, C82.96, C82.97, C82.98, C82.99, C83.00, C83.01, C83.02, C83.03, C83.04, C83.05, C83.06, C83.07, C83.08, C83.09, C83.10, C83.11, C83.12, C83.13, C83.14, C83.15, C83.16, C83.17, C83.18, C83.19, C83.30, C83.31, C83.32, C83.33, C83.34, C83.35, C83.36, C83.37, C83.38, C83.39, C83.50, C83.51, C83.52, C83.53, C83.54, C83.55, C83.56, C83.57, C83.58, C83.59, C83.70, C83.71, C83.72, C83.73, C83.74, C83.75, C83.76, C83.77, C83.78, C83.79, C83.80, C83.81, C83.82, C83.83, C83.84, C83.85, C83.86, C83.87, C83.88, C83.89, C83.90, C83.91, C83.92, C83.93, C83.94, C83.95, C83.96, C83.97, C83.98, C83.99, C84.00, C84.01, C84.02, C84.03, C84.04, C84.05, C84.06, C84.07, C84.08, C84.09, C84.10, C84.11, C84.12, C84.13, C84.14, C84.15, C84.16, C84.17, C84.18, C84.19, C84.40, C84.41, C84.42, C84.43, C84.44, C84.45, C84.46, C84.47, C84.48, C84.49, C84.60, C84.61, C84.62, C84.63, C84.64, C84.65, C84.66, C84.67, C84.68, C84.69, C84.70, C84.71, C84.72, C84.73, C84.74, C84.75, C84.76, C84.77, C84.78, C84.79, C84.90, C84.91, C84.92, C84.93, C84.94, C84.95, C84.96, C84.97, C84.98, C84.99, C84.A0, C84.A1, C84.A2, C84.A3, C84.A4, C84.A5, C84.A6, C84.A7, C84.A8, C84.A9, C84.Z0, C84.Z1, C84.Z2, C84.Z3, C84.Z4, C84.Z5, C84.Z6, C84.Z7, C84.Z8, C84.Z9, C85.10, C85.11, C85.12, C85.13, C85.14, C85.15, C85.16, C85.17, C85.18, C85.19, C85.20, C85.21, C85.22, C85.23, C85.24, C85.25, C85.26, C85.27, C85.28, C85.29, C85.80, C85.81, C85.82, C85.83, C85.84, C85.85, C85.86, C85.87, |

| | |
|---|---|
| | C85.88, C85.89, C85.90, C85.91, C85.92, C85.93, C85.94, C85.95, C85.96, C85.97, C85.98, C85.99, C86.0, C86.1, C86.2, C86.3, C86.4, C86.5, C86.6, C88.0, C88.2, C88.3, C88.4, C88.8, C88.9, C90.00, C90.01, C90.02, C90.10, C90.11, C90.12, C90.20, C90.21, C90.22, C90.30, C90.31, C90.32, C91.00, C91.01, C91.02, C91.10, C91.11, C91.12, C91.30, C91.31, C91.32, C91.40, C91.41, C91.42, C91.50, C91.51, C91.52, C91.60, C91.61, C91.62, C91.90, C91.91, C91.92, C91.A0, C91.A1, C91.A2, C91.Z0, C91.Z1, C91.Z2, C92.00, C92.01, C92.02, C92.10, C92.11, C92.12, C92.20, C92.21, C92.22, C92.30, C92.31, C92.32, C92.40, C92.41, C92.42, C92.50, C92.51, C92.52, C92.60, C92.61, C92.62, C92.90, C92.91, C92.92, C92.A0, C92.A1, C92.A2, C92.Z0, C92.Z1, C92.Z2, C93.00, C93.01, C93.02, C93.10, C93.11, C93.12, C93.30, C93.31, C93.32, C93.90, C93.91, C93.92, C93.Z0, C93.Z1, C93.Z2, C94.00, C94.01, C94.02, C94.20, C94.21, C94.22, C94.30, C94.31, C94.32, C94.40, C94.41, C94.42, C94.6, C94.80, C94.81, C94.82, C95.00, C95.01, C95.02, C95.10, C95.11, C95.12, C95.90, C95.91, C95.92, C96.0, C96.2, C96.20, C96.21, C96.22, C96.29, C96.4, C96.5, C96.6, C96.9, C96.A, C96.Z, D00.00, D00.01, D00.02, D00.03, D00.04, D00.05, D00.06, D00.07, D00.08, D00.1, D00.2, D01.0, D01.1, D01.2, D01.3, D01.40, D01.49, D01.5, D01.7, D01.9, D02.0, D02.1, D02.20, D02.21, D02.22, D02.3, D02.4, D03.0, D03.10, D03.11, D03.111, D03.112, D03.12, D03.121, D03.122, D03.20, D03.21, D03.22, D03.30, D03.39, D03.4, D03.59, D03.60, D03.61, D03.62, D03.70, D03.71, D03.72, D03.8, D03.9, D05.00, D05.01, D05.02, D05.10, D05.11, D05.12, D05.80, D05.81, D05.82, D05.90, D05.91, D05.92, D06.0, D06.1, D06.7, D06.9, D07.0, D07.1, D07.2, D07.30, D07.39, D07.4, D07.5, D07.60, D07.61, D07.69, D09.0, D09.10, D09.19, D09.20, D09.21, D09.22, D09.3, D09.8, D09.9, D18.00, D18.02, D18.03, D18.09, D18.1, D22.0, D22.10, D22.11, D22.111, D22.112, D22.12, D22.121, D22.122, D22.20, D22.21, D22.22, D22.30, D22.39, D22.4, D22.5, D22.60, D22.61, D22.62, D22.70, D22.71, D22.72, D22.9, D25.0, D25.1, D25.2, D25.9, D37.01, D37.02, D37.030, D37.031, D37.032, D37.039, D37.04, D37.05, D37.09, D37.1, D37.2, D37.3, D37.4, D37.5, D37.6, D37.8, D37.9, D38.0, D38.1, D38.2, D38.3, D38.4, D38.5, D38.6, D39.0, D39.10, D39.11, D39.12, D39.2, D39.8, D39.9, D40.0, D40.10, D40.11, D40.12, D40.8, D40.9, D41.00, D41.01, D41.02, D41.10, D41.11, D41.12, D41.20, D41.21, D41.22, D41.3, D41.4, D41.8, D41.9, D42.0, D42.1, D42.9, D43.0, D43.1, D43.2, D43.3, D43.4, D43.8, D43.9, D44.0, D44.10, D44.11, D44.12, D44.2, D44.3, D44.4, D44.5, D44.6, D44.7, D44.9, D45., D46.0, D46.1, D46.20, D46.21, D46.22, D46.4, D46.9, D46.A, D46.B, D46.C, D46.Z, D47.0, D47.01, D47.02, D47.09, D47.1, D47.2, D47.3, D47.4, D47.9, D47.Z1, D47.Z2, D47.Z9, D48.0, D48.1, D48.2, D48.3, D48.4, D48.60, D48.61, D48.62, D48.7, D48.9, D49.0, D49.1, D49.3, D49.4, D49.5, D49.511, D49.512, D49.519, D49.59, D49.6, D49.7, D49.81, D49.89, D49.9, D61.810, D63.0, D64.81, D70.1, E31.22, E31.23, E88.3, G13.1, G73.1, G89.3, H47.42, H47.521, H47.522, H47.529, H47.631, H47.632, H47.639, J91.0, K12.31, K31.7, K63.5, M36.0, M36.1, M84.50XA, M84.511A, M84.512A, M84.519A, M84.521A, M84.522A, M84.529A, M84.531A, |

| | |
|---|---|
| | M84.532A, M84.533A, M84.534A, M84.539A, M84.541A, M84.542A, M84.549A, M84.550A, M84.551A, M84.552A, M84.553A, M84.559A, M84.561A, M84.562A, M84.563A, M84.564A, M84.569A, M84.571A, M84.572A, M84.573A, M84.574A, M84.575A, M84.576A, M84.58XA, N52.36, R18.0, R53.0, T80.810A, T80.810D, T80.810S |
| Cardiomyopathy | ICD9CM: 425, 425.1, 425.11, 425.18, 425.2, 425.3, 425.4, 425.5, 425.7, 425.8, 425.9<br><br>ICD10CM: A36.81, A38.1, A39.50, A39.52, B26.82, B33.20, B33.22, B33.24, B58.81, I40.0, I40.1, I40.8, I40.9, I41., I42.0, I42.1, I42.2, I42.3, I42.4, I42.5, I42.6, I42.8, I42.9, I43., I51.4 |
| Cerebrovascular Disease | ICD9CM: 346.6, 346.61, 346.62, 346.63, 430, 431, 432, 432.1, 432.9, 433, 433, 433.01, 433.1, 433.1, 433.11, 433.2, 433.2, 433.21, 433.3, 433.3, 433.31, 433.8, 433.8, 433.81, 433.9, 433.9, 433.91, 434, 434, 434.01, 434.1, 434.1, 434.11, 434.9, 434.9, 434.91, 435, 435.1, 435.2, 435.3, 435.8, 435.9, 436, 437, 437.1, 437.2, 437.3, 437.4, 437.5, 437.6, 437.7, 437.8, 437.9, 438, 438, 438.1, 438.11, 438.12, 438.13, 438.14, 438.19, 438.2, 438.21, 438.22, 438.3, 438.31, 438.32, 438.4, 438.41, 438.42, 438.5, 438.51, 438.52, 438.53, 438.6, 438.7, 438.81, 438.82, 438.83, 438.84, 438.85, 438.89, 438.9<br><br>ICD10CM: I60.00, I60.01, I60.02, I60.10, I60.11, I60.12, I60.2, I60.20, I60.21, I60.22, I60.30, I60.31, I60.32, I60.4, I60.50, I60.51, I60.52, I60.6, I60.7, I60.8, I60.9, I61.0, I61.1, I61.2, I61.3, I61.4, I61.5, I61.6, I61.8, I61.9, I62.00, I62.01, I62.02, I62.03, I62.1, I62.9, I67.1, I67.2, I67.3, I67.5, I67.6, I67.7, I67.81, I67.82, I67.83, I67.841, I67.848, I67.850, I67.858, I67.89, I67.9, I68.0, I68.2, I68.8 |
| Chronic Kidney Disease | ICD9CM: 285.21, 361.04, 364.76, 403.01, 403.1, 403.11, 403.9, 403.91, 404.01, 404.02, 404.03, 404.1, 404.11, 404.12, 404.13, 404.9, 404.91, 404.92, 404.93, 458.21, 585.1, 585.2, 585.3, 585.4, 585.5, 585.9, 753.1, 753.19, 792.5, 996.56, 996.68, 996.73, V45.1, V45.11, V45.12, V56.0, V56.1, V56.2, V56.31, V56.32, V56.8<br><br>ICD10CM: D63.1, E08.22, E09.22, E10.22, E11.22, E13.22, I12.0, I12.9, I13.0, I13.10, I13.11, I13.2, N18.1, N18.2, N18.3, N18.4, N18.5, N18.6, N18.9, R88.0, Z49.01, Z49.02, Z49.31, Z49.32 |
| Chronic Kidney Failure | ICD9CM: 403.01, 403.11, 403.91, 404.02, 404.03, 404.12, 404.13, 404.92, 404.93, 585.5, 585.6<br><br>ICD10CM: I12.0, I13.11, I13.2, N18.6, N99.0 |
| Chronic Lung Disease | ICD9CM: 491.1, 491.2, 491.21, 491.22, 491.8, 491.9, 492.8, 493, 493.01, 493.02, 493.1, 493.11, 493.12, 493.2, 493.21, 493.22, 493.81, 493.82, 493.9, 493.91, 493.92, 496, 506.4, 516.1, 516.31, 516.4, 516.69, 518.1, 518.2, 518.3, 518.6, 714.81, 748.4, 748.5, 748.61, 748.69<br><br>ICD10CM: B44.81, J41.0, J41.1, J41.8, J42., J43.1, J43.2, J43.8, J43.9, J44.0, J44.1, J44.9, J45.20, J45.21, J45.22, J45.30, J45.31, J45.32, J45.40, J45.41, J45.42, J45.50, J45.51, J45.52, J45.901, J45.902, J45.909, J45.990, J45.991, J45.998, J68.4, |

| | |
|---|---|
| | J70.1, J81.1, J82., J84.03, J84.112, J98.3, M05.10, M05.111, M05.112, M05.119, M05.121, M05.122, M05.129, M05.131, M05.132, M05.139, M05.141, M05.142, M05.149, M05.151, M05.152, M05.159, M05.161, M05.162, M05.169, M05.171, M05.172, M05.179, M05.19, M30.1, Q32.2, Q32.3, Q32.4, Q33.0, Q33.1, Q33.2, Q33.3, Q33.4, Q33.5, Q33.6, Q33.8, Q33.9 |
| Chronic Neuromuscular Disease | ICD9CM: 307.22, 322, 322.1, 322.2, 322.9, 330, 330.1, 330.2, 330.3, 330.8, 330.9, 331, 331.1, 331.11, 331.19, 331.2, 331.3, 331.4, 331.5, 331.6, 331.7, 331.81, 331.82, 331.83, 331.89, 331.9, 333.4, 334, 334.1, 334.2, 334.3, 334.4, 334.8, 334.9, 335.21, 336.2, 337.2, 337.21, 337.22, 337.29, 339.2, 339.21, 339.22, 340, 341, 341.1, 341.2, 341.21, 341.22, 341.8, 341.9, 346.71, 348, 348.1, 348.2, 348.3, 348.3, 348.31, 348.39, 348.4, 348.5, 348.8, 348.81, 348.82, 348.89, 348.9, 354.4, 355.71, 356.3, 357.81, 364, 364.01, 364.02, 364.03, 364.04, 364.05, 364.1, 364.11, 364.21, 364.22, 364.23, 364.24, 364.3, 364.41, 364.42, 364.51, 364.52, 364.53, 364.54, 364.55, 364.56, 364.57, 364.59, 364.6, 364.61, 364.62, 364.63, 364.64, 364.7, 364.71, 364.72, 364.73, 364.74, 364.75, 364.76, 364.77, 364.8, 364.81, 364.82, 364.89, 364.9, 377.1, 377.11, 377.12, 377.13, 377.14, 377.15, 377.16, 378.72, 733.7, 773.4, 774.7, 780.03, 948.4<br><br>ICD10CM: A50.44, E75.23, E83.01, E88.42, F95.1, G03.1, G03.2, G10., G11.4, G14., G23.0, G23.1, G23.2, G30.0, G30.1, G30.8, G30.9, G31.01, G31.09, G31.1, G31.2, G31.81, G31.82, G31.83, G31.85, G31.89, G32.0, G32.89, G35., G36.0, G36.8, G36.9, G37.0, G37.1, G37.2, G37.4, G37.5, G37.8, G37.9, G43.711, G43.719, G56.40, G56.41, G56.42, G56.43, G57.70, G57.71, G57.72, G57.73, G60.1, G61.81, G90.3, G90.50, G90.511, G90.512, G90.513, G90.519, G90.521, G90.522, G90.523, G90.529, G90.59, G91.4, G93.0, H47.20, H47.211, H47.212, H47.213, H47.219, H47.22, H47.231, H47.232, H47.233, H47.239, H47.291, H47.292, H47.293, H47.299, H49.40, H49.41, H49.42, H49.43, I62.03, M89.00, M89.011, M89.012, M89.019, M89.021, M89.022, M89.029, M89.031, M89.032, M89.039, M89.041, M89.042, M89.049, M89.051, M89.052, M89.059, M89.061, M89.062, M89.069, M89.071, M89.072, M89.079, M89.08, M89.09, P57.0, P57.8, P57.9, P91.1, R40.3 |
| Chronic Obstructive Pulmonary Disease | ICD9CM: 115, 115.01, 115.02, 115.03, 115.04, 115.05, 115.09, 115.1, 115.11, 115.12, 115.13, 115.14, 115.15, 115.19, 115.9, 115.91, 115.92, 115.93, 115.94, 115.95, 115.99, 490, 491.1, 491.2, 491.21, 491.22, 491.8, 491.9, 492.8, 494, 494.1, 496, 748.61<br><br>ICD10CM: J41.0 J41.1, J41.8, J42., J43.0, J43.1, J43.2, J43.8, J43.9, J44.0, J44.1, J44.9, J47.0, J47.1, J47.9 |
| Coronary Artery Disease / Heart Disease | ICD9CM: 391.1, 391.2, 391.8, 391.9, 411.8, 411.89, 413.9, 414.01, 414.06, 414.3, 414.4, 414.8, 414.9, 425.1, 425.11, 425.18, 425.2, 425.7, 425.8, 425.9, 440.1, 440.2, 440.21, 440.22, 440.23, 440.24, 440.29, 440.8, 440.9<br><br>ICD10CM: I20.0, I20.1, I20.8, I20.9, I24.0, I24.8, I24.9, I25.10, I25.110, I25.111, I25.118, I25.119, I25.2, I25.5, I25.6, I25.700, |

| | |
|---|---|
| | I25.701, I25.708, I25.709, I25.710, I25.711, I25.718, I25.719, I25.720, I25.721, I25.728, I25.729, I25.730, I25.731, I25.738, I25.739, I25.750, I25.751, I25.758, I25.759, I25.760, I25.761, I25.768, I25.769, I25.790, I25.791, I25.798, I25.799, I25.810, I25.811, I25.812, I25.82, I25.83, I25.84, I25.89, I25.9, Z95.1, Z95.5, Z98.61 |
| Congestive Heart Failure | ICD9CM: 428<br><br>ICD10CM: I50.20, I50.21, I50.22, I50.23, I50.30, I50.31, I50.32, I50.33, I50.40, I50.41 |
| Diabetes Mellitus | ICD9CM: 249.00, 249.01, 249.10, 249.11, 249.20, 249.21, 249.30, 249.31, 249.40, 249.41, 249.50, 249.51, 249.60, 249.61, 249.70, 249.71, 249.80, 249.81, 249.90, 249.91, 250.00, 250.01, 250.02, 250.03, 250.10, 250.11, 250.12, 250.13, 250.20, 250.21, 250.22, 250.23, 250.30, 250.31, 250.32, 250.33, 250.40, 250.41, 250.42, 250.43, 250.50, 250.51, 250.52, 250.53, 250.60, 250.61, 250.62, 250.63, 250.70, 250.71, 250.72, 250.73, 250.80, 250.81, 250.82, 250.83, 250.90, 250.91, 250.92, 250.93, 357.2, 362.01, 362.02, 362.03, 362.04, 362.05, 362.06, 362.07, 366.41, V45.85, V53.91<br><br>ICD10CM: E08.00, E08.01, E08.10, E08.11, E08.21, E08.22, E08.29, E08.311, E08.319, E08.3211, E08.3212, E08.3213, E08.3219, E08.3291, E08.3292, E08.3293, E08.3299, E08.3311, E08.3312, E08.3313, E08.3319, E08.3391, E08.3392, E08.3393, E08.3399, E08.3411, E08.3412, E08.3413, E08.3419, E08.3491, E08.3492, E08.3493, E08.3499, E08.3511, E08.3512, E08.3513, E08.3519, E08.3521, E08.3522, E08.3523, E08.3529, E08.3531, E08.3532, E08.3533, E08.3539, E08.3541, E08.3542, E08.3543, E08.3549, E08.3551, E08.3552, E08.3553, E08.3559, E08.3591, E08.3592, E08.3593, E08.3599, E08.36, E08.37X1, E08.37X2, E08.37X3, E08.37X9, E08.39, E08.40, E08.41, E08.42, E08.43, E08.44, E08.49, E08.51, E08.52, E08.59, E08.610, E08.618, E08.620, E08.621, E08.622, E08.628, E08.630, E08.638, E08.641, E08.649, E08.65, E08.69, E08.8, E08.9, E09.00, E09.01, E09.10, E09.11, E09.21, E09.22, E09.29, E09.311, E09.319, E09.3211, E09.3212, E09.3213, E09.3219, E09.3291, E09.3292, E09.3293, E09.3299, E09.3311, E09.3312, E09.3313, E09.3319, E09.3391, E09.3392, E09.3393, E09.3399, E09.3411, E09.3412, E09.3413, E09.3419, E09.3491, E09.3492, E09.3493, E09.3499, E09.3511, E09.3512, E09.3513, E09.3519, E09.3521, E09.3522, E09.3523, E09.3529, E09.3531, E09.3532, E09.3533, E09.3539, E09.3541, E09.3542, E09.3543, E09.3549, E09.3551, E09.3552, E09.3553, E09.3559, E09.3591, E09.3592, E09.3593, E09.3599, E09.36, E09.37X1, E09.37X2, E09.37X3, E09.37X9, E09.39, E09.40, E09.41, E09.42, E09.43, E09.44, E09.49, E09.51, E09.52, E09.59, E09.610, E09.618, E09.620, E09.621, E09.622, E09.628, E09.630, E09.638, E09.641, E09.649, E09.65, E09.69, E09.8, E09.9, E10.10, E10.11, E10.21, E10.22, E10.29, E10.311, E10.319, E10.321, E10.3211, E10.3212, E10.3213, E10.3219, E10.329, E10.3291, E10.3292, E10.3293, E10.3299, E10.331, E10.3311, E10.3312, E10.3313, E10.3319, E10.339, E10.3391, E10.3392, E10.3393, E10.3399, E10.341, E10.3411, E10.3412, E10.3413, E10.3419, E10.349, E10.3491, E10.3492, E10.3493, E10.3499, E10.351, E10.3511, E10.3512, |

| | |
|---|---|
| | E10.3513, E10.3519, E10.3521, E10.3522, E10.3523, E10.3529, E10.3531, E10.3532, E10.3533, E10.3539, E10.3541, E10.3542, E10.3543, E10.3549, E10.3551, E10.3552, E10.3553, E10.3559, E10.359, E10.3591, E10.3592, E10.3593, E10.3599, E10.36, E10.37X1, E10.37X2, E10.37X3, E10.37X9, E10.39, E10.40, E10.41, E10.42, E10.43, E10.44, E10.49, E10.51, E10.52, E10.59, E10.610, E10.618, E10.620, E10.621, E10.622, E10.628, E10.630, E10.638, E10.640, E10.641, E10.649, E10.65, E10.69, E10.8, E10.9, E11.00, E11.01, E11.10, E11.11, E11.21, E11.22, E11.29, E11.311, E11.319, E11.321, E11.3211, E11.3212, E11.3213, E11.3219, E11.329, E11.3291, E11.3292, E11.3293, E11.3299, E11.331, E11.3311, E11.3312, E11.3313, E11.3319, E11.339, E11.3391, E11.3392, E11.3393, E11.3399, E11.341, E11.3411, E11.3412, E11.3413, E11.3419, E11.349, E11.3491, E11.3492, E11.3493, E11.3499, E11.351, E11.3511, E11.3512, E11.3513, E11.3519, E11.3521, E11.3522, E11.3523, E11.3529, E11.3531, E11.3532, E11.3533, E11.3539, E11.3541, E11.3542, E11.3543, E11.3549, E11.3551, E11.3552, E11.3553, E11.3559, E11.359, E11.3591, E11.3592, E11.3593, E11.3599, E11.36, E11.37X1, E11.37X2, E11.37X3, E11.37X9, E11.39, E11.40, E11.41, E11.42, E11.43, E11.44, E11.49, E11.51, E11.52, E11.59, E11.610, E11.618, E11.620, E11.621, E11.622, E11.628, E11.630, E11.638, E11.640, E11.641, E11.649, E11.65, E11.69, E11.8, E11.9, E13.00, E13.01, E13.10, E13.11, E13.21, E13.22, E13.29, E13.311, E13.319, E13.321, E13.3211, E13.3212, E13.3213, E13.3219, E13.329, E13.3291, E13.3292, E13.3293, E13.3299, E13.331, E13.3311, E13.3312, E13.3313, E13.3319, E13.339, E13.3391, E13.3392, E13.3393, E13.3399, E13.341, E13.3411, E13.3412, E13.3413, E13.3419, E13.349, E13.3491, E13.3492, E13.3493, E13.3499, E13.351, E13.3511, E13.3512, E13.3513, E13.3519, E13.3521, E13.3522, E13.3523, E13.3529, E13.3531, E13.3532, E13.3533, E13.3539, E13.3541, E13.3542, E13.3543, E13.3549, E13.3551, E13.3552, E13.3553, E13.3559, E13.359, E13.3591, E13.3592, E13.3593, E13.3599, E13.36, E13.37X1, E13.37X2, E13.37X3, E13.37X9, E13.39, E13.40, E13.41, E13.42, E13.43, E13.44, E13.49, E13.51, E13.52, E13.59, E13.610, E13.618, E13.620, E13.621, E13.622, E13.628, E13.630, E13.638, E13.640, E13.641, E13.649, E13.65, E13.69, E13.8, E13.9, Z46.81, Z96.41 |
| Drug Dependency | ICD9CM: 304.103, 304.104, 304.105, 304.106, 304.107, 304.108, 304.109, 304.11, 304.11, 304.111, 304.112, 304.113, 304.114, 304.115, 304.116, 304.117, 304.118, 304.119, 304.12, 304.12, 304.121, 304.122, 304.123, 304.124, 304.125, 304.126, 304.127, 304.128, 304.129, 304.13, 304.13, 304.131, 304.132, 304.133, 304.134, 304.135, 304.136, 304.137, 304.138, 304.139, 304.14, 304.15, 304.16, 304.17, 304.18, 304.19, 304.2, 304.21, 304.22, 304.23, 304.3, 304.3, 304.309, 304.31, 304.31, 304.319, 304.32, 304.32, 304.329, 304.33, 304.33, 304.339, 304.39, 304.4, 304.4, 304.401, 304.409, 304.41, 304.41, 304.411, 304.419, 304.42, 304.42, 304.421, 304.429, 304.43, 304.43, 304.431, 304.439, 304.49, 304.5, 304.5, 304.509, 304.51, 304.51, 304.519, 304.52, 304.52, 304.529, 304.53, 304.53, 304.539, 304.59, 304.6, 304.6, 304.609, 304.61, 304.61, 304.619, 304.62, 304.62, 304.629, 304.63, 304.63, 304.639, 304.7, 304.71, 304.72, 304.73, 304.8, 304.81, 304.82, 304.83, |

| | |
|---|---|
| | 304.9, 304.9, 304.909, 304.91, 304.91, 304.919, 304.92, 304.92, 304.929, 304.93, 304.93, 304.939, 304.99<br><br>ICD10CM: F11.20, F11.21, F11.220, F11.221, F11.222, F11.229, F11.23, F11.24, F11.250, F11.251, F11.259, F11.281, F11.282, F11.288, F11.29, F12.20, F12.21, F12.220, F12.221, F12.222, F12.229, F12.250, F12.251, F12.259, F12.280, F12.288, F12.29, F14.20, F14.21, F14.220, F14.221, F14.222, F14.229, F14.23, F14.24, F14.250, F14.251, F14.259, F14.280, F14.281, F14.282, F14.288, F14.29, F15.20, F15.21, F15.220, F15.221, F15.222, F15.229, F15.23, F15.24, F15.250, F15.251, F15.259, F15.280, F15.281, F15.282, F15.288, F15.29, F15.90, F15.920, F15.921, F15.922, F15.929, F15.93, F15.94, F15.950, F15.951, F15.959, F15.980, F15.981, F15.982, F15.988, F15.99 |
| Emphysema | ICD9CM: 492, 492.8, 518.1, 518.2, 770.2, 958.7, 998.81<br><br>ICD10CM: J43.0, J43.1, J43.2, J43.8, J43.9, J98.2, J98.3, P25.0, P25.8, T79.7XXA, T79.7XXD, T79.7XXS, T81.82XA, T81.82XD, T81.82XS |
| Heart Disease | ICD9CM: 391.1, 391.2, 391.8, 391.9, 392, 392.9, 393, 394.1, 395.1, 395.2, 395.9, 397.1, 397.9, 398, 398.9, 398.91, 398.99, 401.1, 401.9, 402, 402.01, 402.1, 402.11, 402.9, 402.91, 404.01, 404.02, 404.03, 404.1, 404.11, 404.12, 404.13, 404.9, 404.91, 404.92, 404.93, 405.01, 405.09, 405.11, 405.19, 405.91, 405.99, 414.01, 414.1, 414.11, 414.12, 414.19, 414.3, 414.4, 415, 415.1, 415.11, 415.12, 415.13, 415.19, 416, 416.1, 416.2, 416.8, 416.9, 417, 417.1, 417.8, 417.9, 424.2, 424.3, 428.1, 428.21, 428.22, 428.23, 428.31, 428.32, 428.33, 428.41, 428.42, 428.43, 428.9, 437.3, 440.1, 440.2, 440.21, 440.22, 440.23, 440.29, 440.8, 441.1, 441.2, 441.3, 441.4, 441.5, 441.6, 441.7, 441.9, 442, 442.1, 442.2, 442.3, 442.81, 442.82, 442.83, 442.84, 442.89, 442.9, 459.3, 459.31, 459.32, 459.33, 459.39, 517.1, 714.4, 725, 796.2, V12.55<br><br>ICD10CM: I00, I01.0, I01.1, I01.2, I01.8, I01.9, I02.0, I02.9, I05.0, I05.1, I05.2, I05.8, I05.9, I06.0, I06.1, I06.2, I06.8, I06.9, I07.0, I07.1, I07.2, I07.8, I07.9, I08.0, I08.1, I08.2, I08.3, I08.8, I08.9, I09.0, I09.1, I09.2, I09.81, I09.89, I09.9, I11.0, I13.0, I13.2, I20.0, I20.1, I20.8, I20.9, I24.0, I24.8, I24.9, I25.10, I25.110, I25.111, I25.118, I25.119, I25.2, I25.3, I25.41, I25.5, I25.6, I25.700, I25.701, I25.708, I25.709, I25.710, I25.711, I25.718, I25.719, I25.720, I25.721, I25.728, I25.729, I25.730, I25.731, I25.738, I25.739, I25.750, I25.751, I25.758, I25.759, I25.760, I25.761, I25.768, I25.769, I25.790, I25.791, I25.798, I25.799, I25.810, I25.811, I25.812, I25.82, I25.83, I25.84, I25.89, I25.9, I27.0, I27.1, I27.2, I27.20, I27.21, I27.22, I27.23, I27.24, I27.29, I27.81, I27.82, I27.83, I27.89, I27.9, I28.0, I28.1, I28.8, I28.9, I50.1, I50.20, I50.21, I50.22, I50.23, I50.30, I50.31, I50.32, I50.33, I50.40, I50.41, I50.42, I50.43, I50.810, I50.811, I50.812, I50.813, I50.814, I50.82, I50.83, I50.84, I50.89, I50.9, I51.0, I51.1, I51.2, I51.3, I51.5, I51.7, I51.81, I51.89, I51.9, I52., I97.130, I97.131, M05.30, M05.311, M05.312, M05.319, M05.321, M05.322, M05.329, M05.331, M05.332, M05.339, M05.341, M05.342, M05.349, M05.351, M05.352, M05.359, M05.361, M05.362, M05.369, M05.371, M05.372, M05.379, M05.39, O29.121, |

| | |
|---|---|
| | O29.122, O29.123, O29.129, Z95.1, Z95.5, Z95.811, Z95.812, Z98.61 |
| Heart Failure (non-congestive) | ICD9CM: 410.52, 410.6, 410.6, 410.61, 410.62, 410.7, 410.7, 410.71, 410.72, 410.8, 410.8, 410.81, 410.82, 410.9, 410.9, 410.91, 410.92, 411, 411.1, 411.8, 411.81, 411.89, 412, 414, 414, 414.01, 414.02, 414.03, 414.04, 414.05, 414.06, 414.07, 414.2, 414.3, 414.4<br><br>ICD10CM: I09.81, I11.0, I13.0, I13.2, I50.1, I50.20, I50.21, I50.22, I50.23, I50.30, I50.31, I50.32, I50.33, I50.40, I50.41, I50.42, I50.43, I50.810, I50.811, I50.812, I50.813, I50.814, I50.82, I50.83, I50.84, I50.89, I50.9, I97.130, I97.131, O29.121, O29.122, O29.123, O29.129, Z95.811, Z95.812 |
| HIV | ICD9CM: 42, 42, 42.1, 42.2, 42.9<br><br>ICD10CM: B20., Z21. |
| Hypertension | ICD9CM: 401, 401.1, 401.9, 402, 402.01, 402.1, 402.11, 402.9, 402.91, 403, 403, 403.01, 403.1, 403.1, 403.11, 403.9, 403.9, 403.91, 404, 404, 404.01, 404.02, 404.03, 404.1, 404.1, 404.11, 404.12, 404.13, 404.9, 404.9, 404.91, 404.92, 404.93, 405.01, 405.09, 405.11, 405.19, 405.91, 405.99, 437.2, 642, 642.01, 642.02, 642.03, 642.04, 642.1, 642.11, 642.12, 642.13, 642.14, 642.2, 642.21, 642.22, 642.23, 642.24, 642.3, 642.31, 642.32, 642.33, 642.34, 642.4, 642.41, 642.42, 642.43, 642.44, 642.5, 642.51, 642.52, 642.53, 642.54, 642.6, 642.61, 642.62, 642.63, 642.64, 642.7, 642.71, 642.72, 642.73, 642.74, 642.9, 642.91, 642.92, 642.93, 642.94<br><br>ICD10CM: I10., O10.011, O10.012, O10.013, O10.019, O10.02, O10.03, O10.911, O10.912, O10.913, O10.919, O10.92, O10.93, O16.1, O16.2, O16.3, O16.4, O16.5, O16.9 |
| Lower Respiratory Infection | ICD9CM: J06.9, J22., J47.0, J80., J98.8<br><br>ICD10CM: 494.1, 518.82, 519.8 |
| Obstructive Sleep Apnea | ICD9CM: 327.23<br><br>ICD10CM: G47.30, G47.31, G47.32, G47.33, G47.34, G47.35, G47.36, G47.37, G47.39 |

**Mechanical Ventilation Outcome,**

| | |
|---|---|
| Mechanical Ventilation | ICD9CM: 96.70, 96.71, 96.72<br><br>ICD10CM: 09HN7BZ, 09HN8BZ, 0DL57DZ, 0DL58DZ, 5A09357, 5A09358, 5A09359, 5A0935B, 5A0935Z, 5A09457, 5A09458, 5A09459, 5A0945B, 5A0945Z, 5A09557, 5A09558, 5A09559, 5A0955B, 5A0955Z, 5A19054, 5A1935Z, 5A1945Z, 5A1955Z,<br><br>CPT4: 4168F, E0463, 94002, 94003, 94004, 94656, 94657 |

**In addition to the ICD-9, ICD-10, and CPT codes listed here, covariates were also defined in the COVID-19 Shared Data Resource domain using natural language processing and keyword text searching of medical encounter notes, laboratory tests, and medications (see text).*

**Supplemental Table 2.** COVID-19 symptoms in the 30 days prior to index date, stratified by PPI user status, unweighted and weighted cohorts

| | UNWEIGHTED COHORT of veterans with positive SARS-CoV-2 test | | WEIGHTED COHORT of veterans with positive SARS-CoV-2 test | | |
|---|---|---|---|---|---|
| **Symptom, N (%)** | PPI non-users (N=8,696) | Current PPI users (N=6,262) | PPI non-users (N=8,696) | Current PPI users (N=6,262) | SMD |
| Asymptomatic | 4,089 (47.0) | 2,644 (42.2) | 4,051 (46.6) | 2,669 (42.6) | 0.080 |
| Chills | 128 (1.5) | 102 (1.6) | 127 (1.5) | 105 (1.7) | 0.017 |
| Cold | 2,430 (27.9) | 1,786 (28.5) | 2,397 (27.6) | 1,800 (28.8) | 0.026 |
| Cough | 2,678 (30.8) | 2,014 (32.2) | 2,687 (30.9) | 2,005 (32.0) | 0.024 |
| Dyspnea | 2,460 (28.3) | 1,996 (31.9) | 2,525 (29.0) | 1,919 (30.6) | 0.035 |
| Fatigue | 507 (5.8) | 463 (7.4) | 539 (6.2) | 428 (6.8) | 0.025 |
| Fever | 2,513 (28.9) | 1,942 (31.0) | 2,513 (28.9) | 1,966 (31.4) | 0.055 |
| Headache | 1,146 (13.2) | 878 (14.0) | 1,125 (12.9) | 897 (14.3) | 0.040 |
| Loss of Smell | 420 (4.8) | 290 (4.6) | 394 (4.5) | 296 (4.7) | 0.009 |
| Loss of Taste | 496 (5.7) | 348 (5.6) | 482 (5.5) | 353 (5.6) | 0.004 |
| Myalgias | 144 (1.7) | 107 (1.7) | 139 (1.6) | 111 (1.8) | 0.014 |
| Rhinorrhea | 11 (0.1) | 5 (0.1) | 11 (0.1) | 5 (0.1) | 0.014 |
| Abdominal Pain | 284 (3.3) | 298 (4.8) | 295 (3.4) | 309 (4.9) | 0.077 |
| Diarrhea | 1,004 (11.5) | 803 (12.8) | 1,017 (11.7) | 802 (12.8) | 0.034 |
| Nausea/Vomiting | 724 (8.3) | 629 (10.0) | 737 (8.5) | 641 (10.2) | 0.061 |

**Supplemental Table 3**. Characteristics of Veterans with SARS-CoV-2 testing (cohort for exploratory analysis), stratified by current PPI user vs. PPI non-user status

| | UNWEIGHTED COHORT of Veterans with SARS-CoV-2 testing | | WEIGHTED COHORT of Veterans with SARS-CoV-2 testing | | |
|---|---|---|---|---|---|
| **COVARIATES** | PPI non-users, n=53,475 | Current PPI users, n=44,199 | PPI non-users, n=53,475 | Current PPI users, n=44,199 | SMD |
| **VHA Facility^** | ^ | ^ | ^ | ^ | 0.027 |
| **Age,** mean years (SD) | 61.1 (15.1) | 64.6 (12.8) | 62.7 (14.3) | 62.8 (14.0) | 0.011 |
| **Male sex, n (%)** | 45,065 (84.3) | 38,841 (12.8) | 45,985 (86) | 38,115 (86.2) | 0.007 |
| **Race/ethnicity, n (%)** | | | | | 0.009 |
| Non-Hispanic White | 28,990 (54.2) | 27,901 (63.1) | 31,049.0 (58.1) | 25,802 (58.4) | |
| Non-Hispanic Black | 13,614 (25.5) | 9,133 (20.7) | 12,536 (23.4) | 10,313 (23.3) | |
| Non-Hispanic other or unknown | 5,008 (9.4) | 3,335 (7.5) | 4,544 (8.5) | 3,360 (8.3) | |
| Hispanic | 5,863 (11.0) | 3,830 (8.7) | 5,346 (10.0) | 4424 (10.0) | |
| **Days from Jan 1 2020 to index date,** mean (SD)[#] | 247 (79.1) | 244 (78.8) | 246 (78.9) | 246 (78.8) | 0.003 |
| **Smoking Status, n (%)** | | | | | 0.044 |
| Current Smoker | 10,725 (20.1) | 8,625 (19.5) | 10,632 (19.9) | 8,797 (19.9) | |
| Former Smoker | 20,673 (38.7) | 20,394 (46.1) | 22,491 (42.1) | 18,811 (42.6) | |
| Never Smoker | 18,212 (34.1) | 13,993 (31.7) | 17,615 (32.9) | 14,735 (33.3) | |
| Unknown | 3,865 (7.2) | 1,187 (2.7) | 2,737 (5.1) | 1,856 (4.2) | |
| **Comorbidities n (%)** | | | | | |
| Asthma | 3,990 (7.5) | 4,679 (10.6) | 4,760.0 (8.9) | 3,975 (9.0) | 0.003 |
| Coronary Artery Disease | 10,600 (19.8) | 13,465 (30.5) | 13,194 (24.7) | 11,021 (24.9) | 0.006 |
| Cancer | 13,471 (25.2) | 14,445 (32.7) | 15,320 (28.6) | 12,808 (29.0) | 0.007 |
| Cardiomyopathy | 1,921 (3.6) | 2,237 (5.1) | 2,312 (4.3) | 1,925 (4.4) | 0.002 |
| Charleson Comorbidity Index, mean (SD) | 1.98 (2.32) | 2.71 (2.58) | 2.32 (2.47) | 2.35 (2.47) | 0.014 |
| Congestive Heart Failure | 4,339 (8.1) | 5,578 (12.6) | 5,456 (10.2) | 4,543 (10.3) | 0.002 |
| Chronic Lung Disease | 17,911 (33.5) | 20,876 (47.2) | 21,307 (39.8) | 17,814 (40.3) | 0.009 |
| Chronic Neuromuscular Disease | 2,126 (4.0) | 2,133 (4.8) | 2,357 (4.4) | 1,971 (4.5) | 0.003 |
| Chronic Kidney Disease | 7,488 (14.0) | 8,382 (19.0) | 8,755 (16.4) | 7,301 (16.5) | 0.004 |
| Chronic Kidney Failure | 1,005 (1.9) | 1,031 (2.3) | 1,126 (2.1) | 942 (2.1) | 0.002 |
| Chronic Obstructive Pulmonary Disease | 9,819 (18.4) | 13,103 (29.6) | 12,589 (23.5) | 10,526 (23.8) | 0.006 |
| Central Sleep Apnea | 206 (0.4) | 249 (0.6) | 257 (0.5) | 214 (0.5) | <0.001 |
| Cardiovascular Disease | 18,936 (35.4) | 21,655 (49.0) | 22,314 (41.7) | 18,621 (42.1) | 0.008 |
| Diabetes | 17,393 (32.5) | 18,799 (42.5) | 19,867 (37.2) | 16,551 (37.4) | 0.006 |
| Drug Dependency | 4,094 (7.7) | 3,390 (7.7) | 4,076 (7.6) | 3,391 (7.7) | 0.002 |
| Emphysema | 1,053 (2.0) | 1,461 (3.3) | 1,369 (2.6) | 1,153 (2.6) | 0.003 |
| Heart Disease | 13,506 (25.3) | 16,318 (36.9) | 16,388 (30.6) | 13,650 (30.9) | 0.005 |
| Heart Failure (non-congestive) | 5,381 (10.1) | 6,799 (15.4) | 6,705 (12.5) | 5,572 (12.6) | 0.002 |
| *H. pylori* positive | 10,114 (18.9) | 7,217 (16.3) | 9491 (17.7) | 7,813 (17.7) | 0.005 |
| HIV | 846 (1.6) | 407 (0.9) | 706 (1.3) | 602 (1.4) | 0.004 |
| Hypertension | 31,542 (59.0) | 32,847 (74.3) | 35,307 (66.0) | 29,548 (66.9) | 0.018 |
| Lower Respiratory Infection | 5,570 (10.4) | 5,468 (12.4) | 6,082 (11.4) | 5,074 (11.5) | 0.003 |
| Obstructive Sleep Apnea | 16,711 (31.3) | 18,792 (42.5) | 19,519 (36.5) | 16,402 (37.1) | 0.014 |
| **Medications, n (%)** | | | | | |
| ACE Inhibitors | 14,334 (26.8) | 15,559 (35.2) | 16,472 (30.8) | 13,777 (31.2) | 0.008 |
| ARBs | 6,808 (12.7) | 8,221 (18.6) | 8,240 (15.4) | 6,923 (15.7) | 0.007 |
| H2RAs | 3,902 (7.3) | 3,587 (8.1) | 4,246 (7.9) | 3,587 (8.1) | 0.006 |

| | | | | | |
|---|---|---|---|---|---|
| NSAIDs | 34,486 (64.5) | 34,028 (77.0) | 37,541 (70.2) | 31,520 (71.3) | 0.024 |
| Statins | 25,878 (48.4) | 29,841 (67.5) | 30,506 (57.0) | 25,674 (58.1) | 0.021 |

*Abbreviations: Angiotensin converting enzyme inhibitors (ACE inhibitors); Angiotensin II receptor blockers (ARBs); Helicobacter pylori (H. pylori); Histamine-2 receptor antagonists (H2RAs); Non-steroidal anti-inflammatory agents (NSAIDs); Proton pump inhibitor (PPI); standard deviation (SD); standardized mean difference (SMD); Veterans Health Administration (VHA)*

*^Each of the 127 VHA facilities were included as covariates in this analysis; however, the proportion of patients at each station for each group is not listed here due to space considerations.*

*[#]This variable represents the days from January 1, 2020 to the index date of SARS-CoV-2 testing to account for temporal differences. Please refer text for additional details.*

**Supplemental Figure 1. Proportion of SARS-CoV-2 positive testing among veterans tested through January 9, 2021.** Histogram illustrating the number of SARS-CoV-2 tests performed within the VHA nationwide between March 1, 2020 through January 9, 2020, and the proportion of positive test results. The magnitude and temporality of the SARS-CoV-2 positivity rate reflects the overall observed trend nationally during this time period.

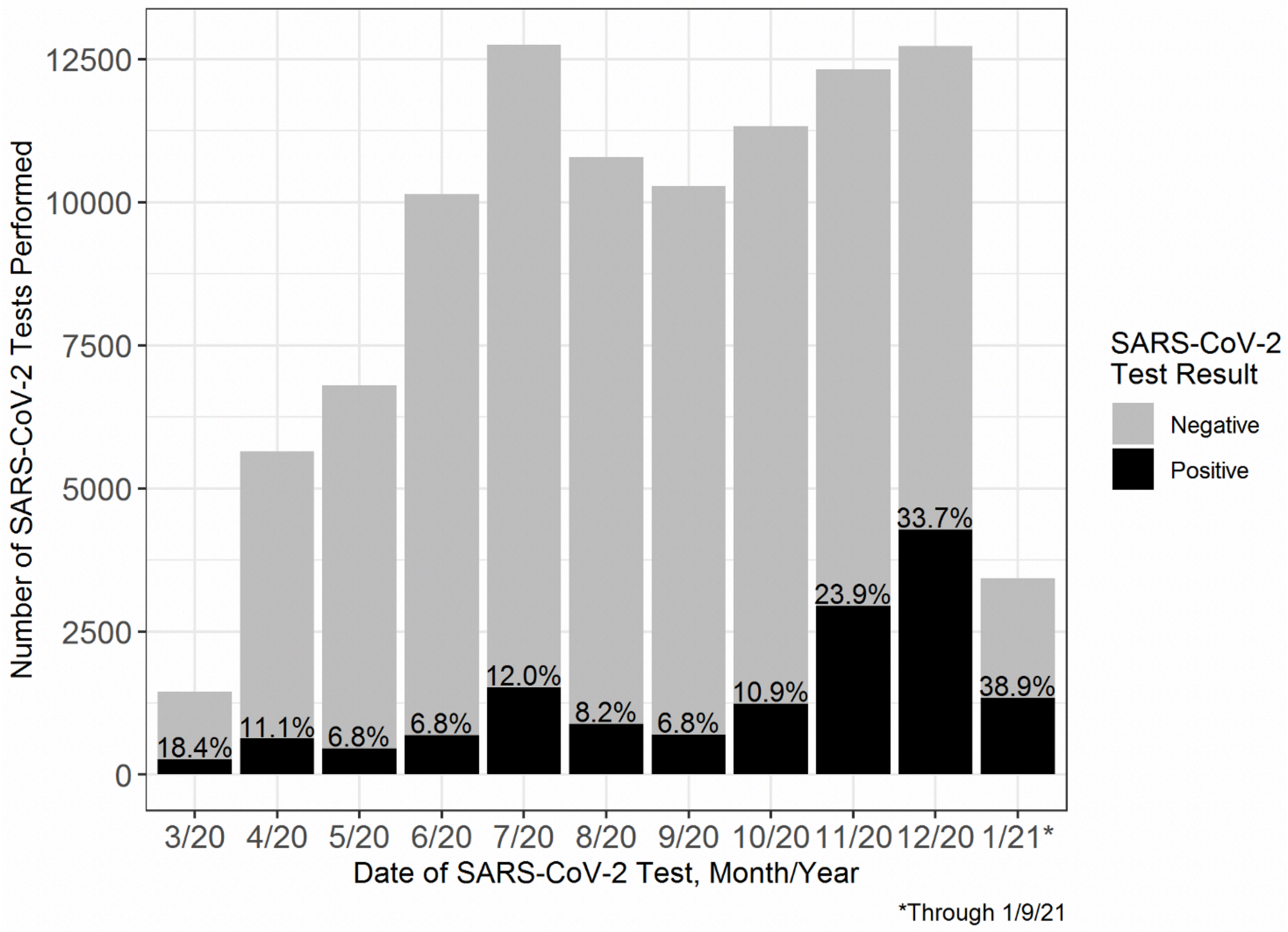

**Supplemental Figure 2. Distribution of propensity scores for current PPI users vs. non-users among veterans who tested positive for SARS-CoV-2 (N=14,958; primary analytic cohort)**. Propensity score (PS) weighting was used to balance covariate distributions between PPI users and PPI non-users. We calculated the Average Treatment Effect (ATE) weights as the inverse of the probability of the observed PPI use statuses. Weights were scaled by a constant so that the sum of weights equaled the unweighted cohort's sample size. To balance the large number of covariates, PS were calculated via cross-validated logistic ridge regression models with optimal shrinkage parameters. To balance nonlinear associations with PPI use, the PS models included restricted cubic splines on all continuous covariates, including the date of positive SARS-CoV-2 testing (index date). As determined based on sufficient overlap in the PS, balance was achieved without extreme weights (see Supplemental Figure 3)

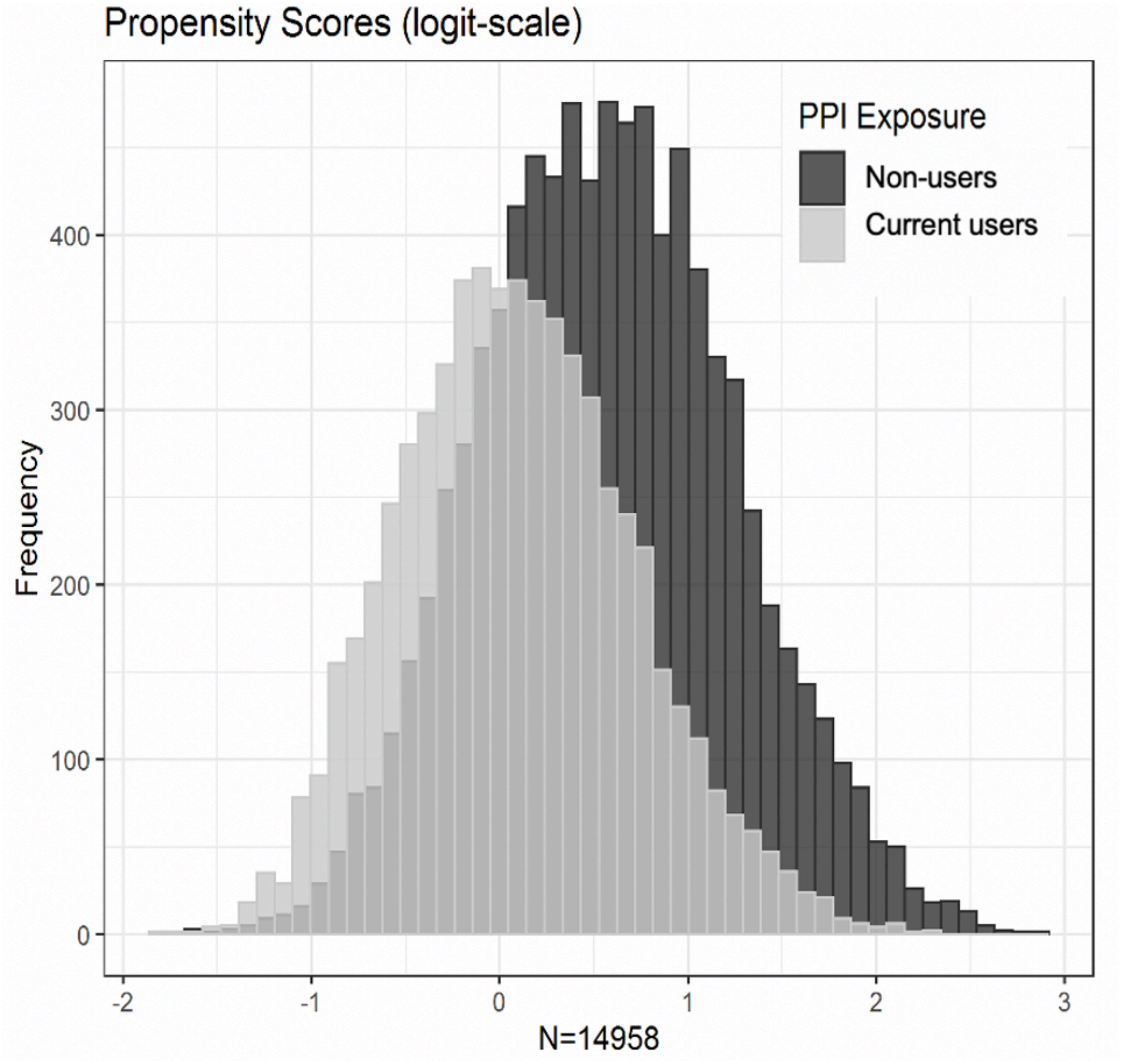

**Supplemental Figure 3. Plot of standardized mean differences for covariates between current PPI users and non-users for the unweighted and weighted cohorts.** Standardized mean differences (SMDs) were used to compare the means and standard deviations (SD) for continuous variables and proportions for categorical variables between current PPI users and non-users. SMDs are the preferred measure of covariate balance in large cohorts. Smaller SMDs indicate better balance between groups, with a threshold of <0.1 (red vertical line) indicating adequate balance. As depicted in the SMD plot of both the unweighted and propensity score-weighted cohort, full covariate balance was achieved in the weighted cohort.

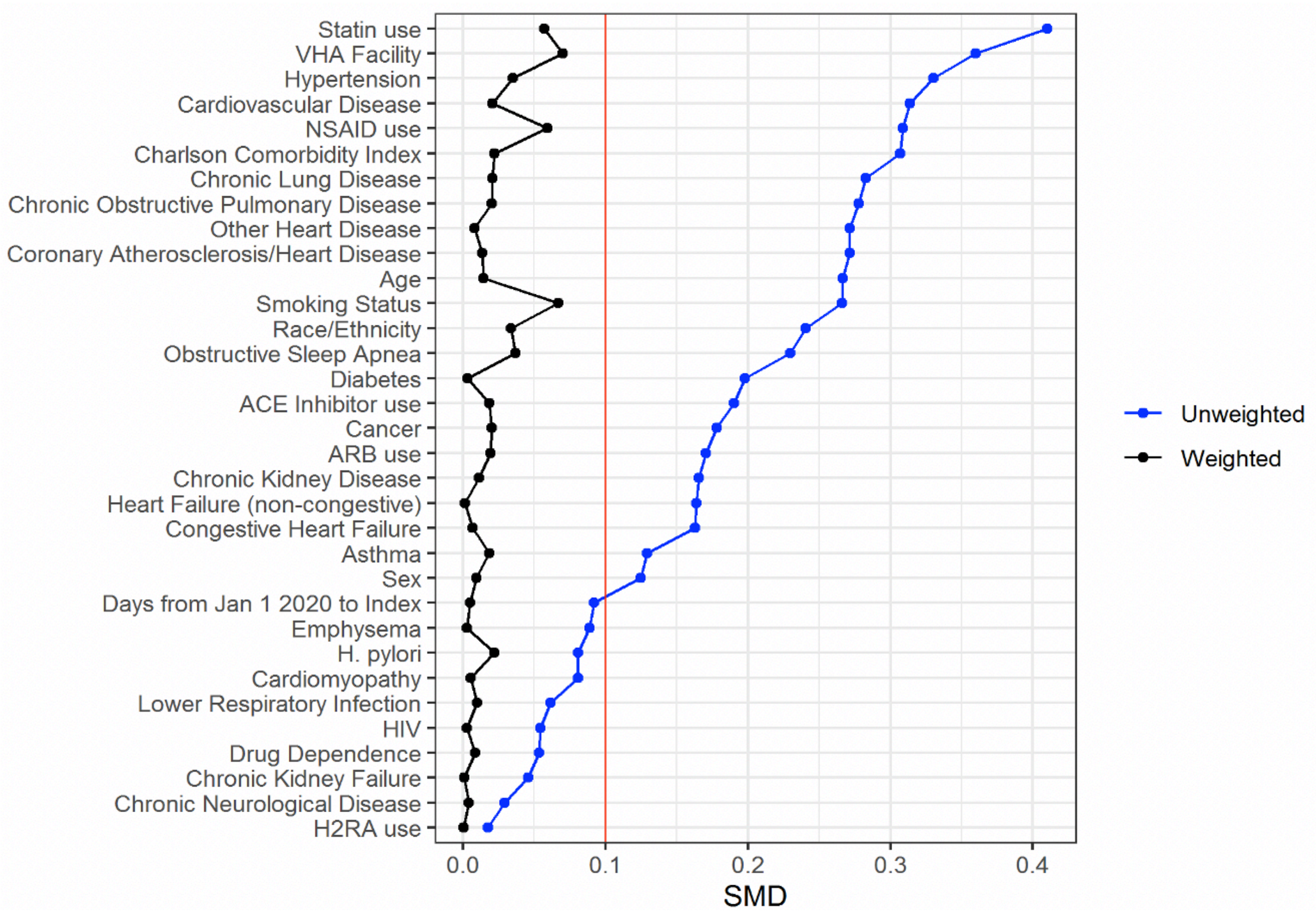
